# Longitudinal Analysis of Superoxide Dismutase 1 Seeding Activity in Amyotrophic Lateral Sclerosis Cerebrospinal Fluid

**DOI:** 10.64898/2026.03.20.26348753

**Authors:** Mylene A. Sebogo, Macie C. Frans, Helan Paulose, Claire L. Rodriguez, Ging-Yuek Hsiung, Neil R. Cashman, Cindy V. Ly, Moses J. Leavens

## Abstract

Twenty percent of familial amyotrophic lateral sclerosis (fALS) cases are linked to mutations in the Superoxide Dismutase 1 (*SOD1)* gene and accumulation of misfolded SOD1 aggregates. SOD1 misfolding from the broader ALS population without *SOD1* mutations is less clear. Here, we report SOD1 seeding activity in antemortem cerebrospinal fluid (CSF) from ALS participants with and without *SOD1* mutations during ALS progression. Antemortem CSF from controls, *SOD1-*ALS, and sporadic ALS (sALS) patients was subjected to SOD1 seed amplification real-time quaking induced conversion (RT-QuIC) assays. *SOD1*-ALS CSF exhibited shorter lag phase and increased ThioflavinT (ThT) fluorescence amplitude compared to healthy controls and those with spinal muscular atrophy. CSF from sALS participants, who had no mutations in *SOD1* or nine other ALS risk genes, also displayed SOD1 seeding activity, indicating wild-type SOD1 is aggregate-prone in the broader ALS population. Longitudinal CSF data indicated that SOD1 seeding activity correlates with ALS progression via the ALS Functional Rating Scale Revised (ALSFRS-R) slope decline and CSF neurofilament light. Our sALS CSF cohort primarily comprised of participants less than 2 years from symptom onset, suggesting that SOD1 seeding activity is an early biomarker that may enable inclusion in clinical trials. With the FDA-approval of tofersen (Qalsody), a SOD1-lowering antisense oligonucleotide, new SOD1 diagnostic, prognostic and pharmacodynamic biomarkers may enable SOD1-targeting strategies that could benefit the broader ALS population.

## Introduction

Amyotrophic Lateral Sclerosis (ALS) is a rapidly progressive neurodegenerative disease caused by motor neuron loss manifesting as limb, respiratory, and bulbar dysfunction. On average, three to five years after receiving an ALS diagnosis, patients succumb to disease due to respiratory failure [1,2]. About 10% of ALS cases is familial (fALS), while the remaining 90% are sporadic ALS (sALS) [1]. The most common genetic mutations linked to ALS are chromosome 9 open reading frame 72 (*C9ORF72*), superoxide dismutase 1 (*SOD1*), transactivation response DNA binding protein of 43 kD (*TARDBP*), and fused in sarcoma (*FUS*) [2]. Currently, ALS has three Food and Drug Administration approved treatments; riluzole and edavarone, which provide modest benefit in sALS, and tofersen (Qalsody), which in 2023 was given accelerated FDA approval for ALS patients with mutations in *SOD1*, approximately 2% of the ALS population [3,4]. Treatment options for many ALS patients remain limited.

Clinical manifestations of ALS, in sALS or ALS linked to genetic mutations, are similar [5], indicating pathological features may be shared between all ALS. Thus, the identification of shared pathological features may uncover new biomarkers and targets for therapeutic intervention across the entire ALS population. ALS is characterized by perturbations in retrotransposon activation [6], RNA metabolism [7], oxidative and proteotoxic stress [5,8–11], disruptions in axon maintenance [12–14], and impairment of mitochondria [15–18]. Transcriptome analyses from postmortem ALS motor cortices suggest molecular classes corresponding to oxidative and proteotoxic stress, activation of glia, and high levels of retrotransposon expression with dysfunction of Transactivation DNA Response Binding protein of 43 kD (TDP-43) [8], underscoring the need to identify and characterize distinct clinical subgroups to refine recruitment for clinical trials and guide targeted therapeutic development. With respect to oxidative and proteotoxic stress, several studies have indicated misfolded SOD1 is a component of *SOD1*-linked and non-*SOD1* linked ALS pathology [19–24], but fluid biomarkers that can measure disease-associated SOD1 with seeding activity from antemortem ALS biospecimens have not been available.

SOD1 is a copper and zinc dismutase that functions as a homodimer to dismutase superoxide to molecular oxygen and hydrogen peroxide [25], an essential antioxidant enzyme [26,27]. Besides dismutase activity, other SOD1 functions have been reported [28]. Mutations in *SOD1* are responsible for 12–20% of all fALS cases, with the p.A5V mutation comprising ∼30% of *SOD1*-ALS cases in North America [29]. Several *SOD1* variants are implicated in ALS, conferring heterogeneity with respect to clinical progression and symptom onset age [29,30], with differences in SOD1 stability and dismutase activity [31–33]. Mutations in *SOD1* confer susceptibility to protein misfolding and aggregation [34–36], in the SOD1 apo state [26,33,37,38] and upon SOD1 oxidization [35,39,40].

Wild-type (WT) SOD1 has also been reported to misfold under oxidative or metal-free conditions, linking aberrant WT SOD1 structure to sALS pathology [19,21,41]. In sALS postmortem tissues, SOD1 inclusions [19] and accumulation of disordered and abnormally post-translationally modified SOD1 species from those with or without genetic mutations have been reported [21][19]. Seeding activity of SOD1 from SOD1-ALS, C9ORF72-ALS, and sALS postmortem motor cortex and spinal cord homogenates have been observed using an SOD1 seed amplification real-time quaking induced conversion (RT-QuIC) assay [41]. Together, these studies indicate non-*SOD1* linked ALS cases may have seeding competent WT SOD1 in the broader sALS population.

Evidence for disease-associated SOD1 aggregates in accessible, antemortem ALS biospecimens has been reported [24], but SOD1 seeding activity is unknown. Here, we used a SOD1 seed amplification RT-QuIC assay to probe for seeding competent WT and mutant SOD1 from ALS participants relatively early in disease (< 2 years from symptom onset) during their progression. We observed the rate of CSF SOD1 seeding activity via slope and ThT fluorescence amplitude correlated with ALS progression and with CSF neurofilament light, highlighting potential new biomarker applications in light of ongoing clinical trials with SOD1 targeted therapies.

## Materials and Methods

### Preparation of human SOD1 plasmid

cDNA encoding human WT SOD1 (rSOD1) was manufactured by Genscript (Piscataway NJ, USA) and placed in the pET 28a(+) vector with an N-terminus 6x His-tag (**Table S1**). SOD1 cDNA sequence was confirmed by Genscript and the mass of rSOD1 was confirmed with electrospray ionization liquid chromatography mass spectrometry (ESI LC-MS).

### Human SOD1 expression and purification

This protocol has been previously described in detail [41]. The SOD1 cDNA was expressed from the pET 28a(+) vector by transformation into BL21 (New England Biolabs, Ipswich MA, USA; DE3, #C2727I) *E. coli* competent cells. A liter of sterile 2xYT media in a 2.8 L Fernbach flask was inoculated with 10 mL of transformed *E. coli* cells and 1 mL of 50 mg/mL kanamycin. Cells were grown in an orbital shaker at 250 rpm and 37 °C, 1 mM final IPTG (GoldBio, #I2481C) was added when OD_600_ was between 0.6 - 0.8, and cells grew for ∼15 hours at 30 °C and 150 rpm. Media was spun for 15 minutes, 7,500 rpm at 4 °C. Using cell pellets from 2 L liters of media, ∼100 mL of 0.5 M NaCl, 0.05 M Tris, pH 8.0 buffer was used with 0.5 mg/mL lysozyme, put on ice on stir plate for 30 minutes, with sonication for 5 minutes (30 seconds on/30 seconds off, 35% AMPL) using a probe sonicator (Sonics VibraCell, Newtown CT, USA). Lysate was spun for 30 minutes at 12,500 rpm and 4 °C, and 50 mL of filtered supernatant was used for affinity chromatography. A Bio-Rad (Hercules CA, USA) NGC fast protein liquid chromatography system with a 5 mL HisTrap HP column (Cytiva, Marlborough MA, USA) was washed with 0.1 M sodium phosphate, 0.3 M NaCl, 0.02 M imidazole, 2 mM β-ME, pH 7.4 buffer (buffer A). Fifty mL of supernatant was applied to the column, washed with buffer A, and eluted with buffer A in 0.3 M imidazole using a 0-100% imidazole gradient. Protein was then buffer exchanged overnight (∼15 hours) at room temperature (∼22 °C) with 25 mM Tris pH 8.0 using 10,000 MWCO Snakeskin dialysis tubing (ThermoScientific, #68100). Next day, dialysate was spun for 30 min, 15,000 rpm at 20 °C. Supernatant was concentrated in a 10,000 MWCO Amicon centrifugal concentrator (Millipore Sigma, Burlington MA, USA), and protein purity was assessed with ESI LC-MS with monomeric mass of 17,866 Da (**Fig. S1**).

### Preparation of Antemortem Cerebrospinal Fluid

Human antemortem cerebrospinal fluid (CSF) from 32 controls (13 disease controls, 19 healthy controls) and from 23 ALS participants clinically diagnosed with *SOD1*-related ALS (n=5) or sporadic ALS (n=18) were obtained from the Target ALS Longitudinal Biofluids core, Dr. Cindy Ly at Washington University, Drs. Neil Cashman and Ging-Yuek Hsiung at University of British Columbia, Innovative Research, and Precision for Medicine (**Table 1**). Thirteen of eighteen sporadic ALS cases had whole exome sequencing to confirm ALS risk mutations in 10 different genes including *SOD1* (**Table 2**). Longitudinal CSF collections were available from 18 of 23 ALS representing samples acquired 13-168 months post-symptom onset for visit 1 and 3-24 months post visit 1 for visit 2. CSF was received on dry ice, aliquoted, and stored at –80 °C.

**Table 1.** Control and ALS participant information.

| Controls | Age range (y) | Gender | Mutation | Diagnosis |
| --- | --- | --- | --- | --- |
| Control 1 | 41-45 | Female | - | - |
| Control 2 | 56-60 | Female | - | - |
| Control 3 | 66-70 | Male | - | - |
| Control 4 | 56-60 | Female | - | - |
| Control 5 | 66-70 | Male | - | - |
| Control 6 | 41-45 | Male | - | - |
| Control 7 | - | - | - | - |
| Control 8 | 56-60 | Female | - | - |
| Control 9 | 51-55 | Female | - | - |
| Control 10 | 41-45 | Female | - | - |
| Control 11 | 46-50 | Female | - | - |
| Control 12 | 36-40 | Male | *C9ORF72* | - |
| Control 13 | 56-60 | Female | - | - |
| Control 14 | 66-70 | Male | - | - |
| Control 15 | 51-55 | Female | *C9ORF72* | - |
| Control 16 | 56-60 | Male | *C9ORF72* | - |
| Control 17 | 31-35 | Female | *SMN1/2* | SMA Type 3 |
| Control 18 | 21-25 | Male | *SMN1/2* | SMA Type 3 |
| Control 19 | 56-60 | Male | *SMN1/2* | SMA Type 3/4 |
| Control 20 | 36-40 | Male | *SMN1/2* | SMA Type 3 |
| Control 21 | 56-60 | Female | - | AD |
| Control 22 | 61-65 | Male | - | PD |
| Control 23 | 55-60 | Female | - | MCI |
| Control 24 | 51-55 | Male | *C9ORF72* | FTD-ALS |
| Control 25 | 46-50 | Male | - | HIV+, subject complaints only |
| Control 26 | 46-50 | Male | - | anxiety, subject complaints only |
| Control 27 | 66-70 | Male | - | anxiety, subject complaints only |
| Control 28 | 66-70 | Male | - | HIV+, subject complaints only |
| Control 29 | 46-50 | Male | - | HIV+, subject complaints only |
| Control 30 | 46-50 | Female | *C9ORF72* | BvFTD |
| Control 31 | 46-50 | Female | *C9ORF72* | BvFTD |
| Control 32 | 61-65 | Male | *C9ORF72* | BvFTD |

| ALS participants | Age range (y) | Gender | Mutation | ALS Diagnosis (El Escorial) | Onset year | Site of onset | Diagnosis year | Disease duration (onset to visit 1, months) | Disease duration (onset to visit 2, months) | Medications |
| --- | --- | --- | --- | --- | --- | --- | --- | --- | --- | --- |
| *SOD1*-ALS 1 | 51-55 | M | p.Val32Ala | Definite | 2013 | Other | 2017 | 96 | 105 | tofersen |
| *SOD1*-ALS 2 | 51-55 | M | p.Glu101Lys | Definite | 2016 | Limb | 2019 | 65 | 81 | tofersen |
| *SOD1*-ALS 3 | 56-60 | M | p.Ala90Val | Definite | 2007 | Limb | 2008 | 168 | 192 | tofersen |
| *SOD1*-ALS 4 | 71-75 | M | p.Leu145Ser | Definite | 2015 | Limb | 2017 | 94 | 103 | Riluzole, tofersen |
| *SOD1*-ALS 5 | 46-50 | M | ° | Possible | 2023 | Bulbar | 2023 | - | - | Riluzole, tofersen |
| sALS 1 | 66-70 | F | None | Probable | 2018 | Limb | 2021 | 60 | 72 | Riluzole |
| sALS 2 | 61-65 | M | None | - | - | Limb | - | 21 | 34.8 | N/A |
| sALS 3 | 66-70 | M | None | Definite | 2019 | Limb | 2020 | 29 | 46 | None |
| sALS 4 | 31-35 | M | None | Definite | 2020 | Limb | 2020 | 30 | 34 | None |
| sALS 5 | 61-65 | M | None | Definite | 2018 | Limb | 2019 | 36 | 39 | Riluzole |
| sALS 6 | 51-55 | F | None | Probable | 2012 | Limb | 2014 | 111 | 127 | Riluzole, edavarone |
| sALS 7 | 51-55 | M | None | Probable | 2017 | Limb | 2018 | 59 | 76 | None |
| sALS 8 | 26-30 | M | None | Probable | 2020 | Limb | 2022 | 18 | 25 | Riluzole, edavarone |
| sALS 9 | 56-60 | F | None | Probable | 2021 | Bulbar | 2021 | 13 | 20 | Riluzole, edavarone |
| sALS 10 | 61-65 | F | None | Probable | 2021 | Limb | 2023 | 16 | 23 | None |
| sALS 11 | 61-65 | F | None | Probable | 2018 | Limb | 2020 | 53 | 56 | Riluzole, edavarone |
| sALS 12 | 66-70 | M | None | Probable | 2022 | Limb | 2022 | 16 | 22 | Riluzole, edavarone |
| sALS 13 | 61-65 | M | None | Probable | 2022 | Limb | 2022 | 17 | 21 | Riluzole, edavarone |
| sALS 14 | 56-60 | F | Unknown | - | - | - | - | - | - | - |
| sALS 15 | 66-70 | F | Unknown | - | - | - | - | - | - | - |
| sALS 16 | 71-75 | M | Unknown | - | - | - | - | - | - | - |
| sALS 17 | 71-75 | F | Unknown | - | - | - | - | - | - | - |
| sALS 18 | 56-60 | F | Unknown | ALS, no FTD | Unknown | Unknown | - | 36 | - | - |

**Table 2.** presence or absence of genetic mutation in ALS participants.

| ALS participant | *SOD1* | *C9ORF72* | *TARDBP* | *FUS* | *MAPT* | *ANG* | *VCP* | *VAPB* | *GRN* | *SETX* |
| --- | --- | --- | --- | --- | --- | --- | --- | --- | --- | --- |
| *SOD1*-ALS 1 | + | - | - | - | - | - | - | - | - | - |
| *SOD1*-ALS 2 | + | - | - | - | - | - | - | - | - | - |
| *SOD1*-ALS 3 | + | - | - | - | - | - | - | - | - | - |
| *SOD1*-ALS 4 | + | - | - | - | - | - | - | - | - | - |
| *SOD1*-ALS 5 | + | - | - | - | - | - | - | - | - | - |
| sALS 1 | - | - | - | - | - | - | - | - | - | - |
| sALS 2 | - | - | - | - | - | - | - | - | - | - |
| sALS 3 | - | - | - | - | - | - | - | - | - | - |
| sALS 4 | - | - | - | - | - | - | - | - | - | - |
| sALS 5 | - | - | - | - | - | - | - | - | - | - |
| sALS 6 | - | - | - | - | - | - | - | - | - | - |
| sALS 7 | - | - | - | - | - | - | - | - | - | - |
| sALS 8 | - | - | - | - | - | - | - | - | - | - |
| sALS 9 | - | - | - | - | - | - | - | - | - | - |
| sALS 10 | - | - | - | - | - | - | - | - | - | - |
| sALS 11 | - | - | - | - | - | - | - | - | - | - |
| sALS 12 | - | - | - | - | - | - | - | - | - | - |
| sALS 13 | - | - | - | - | - | - | - | - | - | - |
| sALS 14 | NE | NE | NE | NE | NE | NE | NE | NE | NE | NE |
| sALS 15 | NE | NE | NE | NE | NE | NE | NE | NE | NE | NE |
| sALS 16 | NE | NE | NE | NE | NE | NE | NE | NE | NE | NE |
| sALS 17 | NE | NE | NE | NE | NE | NE | NE | NE | NE | NE |
| sALS 18 | NE | NE | NE | NE | NE | NE | NE | NE | NE | NE |
| + = presence of mutation, - = absence of mutation, NE = not evaluated. *SOD1* = Superoxide Dismutase 1, *C9ORF72* = Chromosome 9 opening reading frame 72, *TARDBP* = Transactivation response DNA binding protein 43 kD, *FUS* = Fused in sarcoma, *MAPT* = tau, *ANG* = Angiogenin, *VCP* = Valosin-containing protein, *VAPB* = Vesicle-associated membrane associated protein, *GRN* = Progranulin, *SEXT* = Senataxin. |  |  |  |  |  |  |  |  |  |  |

### SOD1 RT-QuIC Assay for Antemortem CSF

After dialysis, substrate was spun, concentrated, and filtered (100 kD Pall centrifugal filter #OD100C35, LOT #FL6611) at 2,000x*g*, 20 °C, for 10 minutes prior to concentrating to a stock concentration of ∼290 µM (ε = 5,500 M^-1^cm^-1^, Thermo Scientific NanoDrop One). SOD1 RT-QuIC buffer solution was prepared with a 100 mL volumetric flask using ≥99% GuHCl (Sigma Aldrich #G3272), 99% sodium acetate (Alfa Aesar #127093), and high-performance liquid chromatography grade ddH_2_O (Sigma Aldrich #270733) at 0.52 M GuHCl, 0.02 M sodium acetate, pH 4.0. Fifteen µM of Thioflavin T (ThT) and 15 µM rSOD1 (∼0.26 mg/mL, 5,500 M^-1^cm^-1^) was used in each 96-well microplate (Thermo-Scientific Nunc 96-well optical bottomed black polystyrene plates w/Lid, catalog #165305). Ninety-five µL of reaction mix (15 µM ThT, 15 µM rSOD1, 0.52 M GuHCl, 0.02 M sodium acetate, pH 4.0) was pipet in each well. 5 µL of ALS or control CSF (diluted 2:1 in ddH_2_O, Sigma Aldrich #270733) was pipet in each well. Reaction mix alone (15 µM ThT, 15 µM rSOD1, SOD1 RT-QuIC buffer) at 95 µL was used in each plate as non-CSF control. Plates were sealed with tape (Thermo-Scientific, clear polyolefin, non-sterile, catalog #232702) and were put in FLUOstar Omega readers (BMG Labtech, Germany) at 37 °C, 400 rpm, with 25 seconds of shaking and 25 seconds of resting on a double orbital setting. ThT fluorescence intensity measurements were collected every 40 minutes with 448 nm excitation and 482 nm emission using a gain setting of 1,500. CSF samples were replicated in quadruplicate; experimenter was blinded to the identity of CSF samples.

### CSF Neurofilament light Simoa immunoassay

CSF Neurofilament light (NfL) concentrations were measured in ALS participant CSF at both visits (n=18, 5 *SOD1*-ALS, 13 sALS) using the NF-Light v2 Advantage kit on a Simoa HDx analyser (Quanterix). The assay was performed as per the manufacturer’s protocol.

### Antibody incubation with CSF to assess specificity of SOD1 RT-QuIC Assay

Addition of anti-SOD1 C4F6 antibody (MediMabs #MM-0070-2-P) and control antibody (ThermoFisher #MA110418) were prepared according to the manufacturer’s instructions at 0.5 µg/µL. Antibodies were incubated with ALS or control CSF for 1 hour at room temperature with final antibody concentration in each well equal to 50 ng/µL. 5 µL of CSF-antibody mix was added to 95 µL reaction mix using CSF SOD1 RT-QuIC assay protocol.

### SOD1 RT-QuIC Statistical Analysis

Based on difference in proportions, we estimated 16 CSF samples from ALS cases and controls would provide 90% power to demonstrate a significant difference in seeding activity (p<0.05). We examined 23 ALS CSF samples and 32 control CSF samples (**Table 1**). After experiments, BMG Omega Mars .RUC files were exported to Microsoft Excel, SigmaPlot 14.5 or 15.0 (Systat software), or GraphPad Prism 10 for data analyses. To compare CSF data from controls and ALS in each plate, we subtracted the final minus the initial reading from each technical replicate, averaged the four technical replicates, then used a Student’s t-test or Mann Whitney test (sALS visit 2 only because normality test failed) to compare the mean or median ThT fluorescence. To estimate lag phase, slope, and ThT fluorescence amplitude, we fit the raw data using equation 1 [41]. In equation 1,

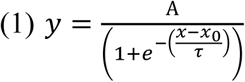

where A is ThT fluorescence amplitude, x_0_ is the time to reach midpoint ThT fluorescence, and τ is a fibril time constant, where lag phase equals x_0_ – 2τ [41,42]. Raw kinetic traces were fit to this model when able using no constraints to estimate x_0,_ τ, and A. We took the first derivative of Eq 1 to determine the slope for each CSF SOD1 RT-QuIC kinetic curve (full derivation in **Table S2**). In equation 2,

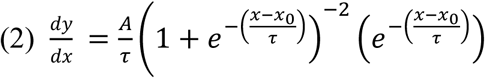

where A, τ, and x_0_ are parameters defined above. To determine slope at the midpoint, x equals x_0_, and equation 2 reduces to equation 3.

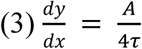

Equation 3 is the rate of CSF SOD1 seeding activity at the midpoint, with relative fluorescence units per hour (RFU/hr). We used A and τ from fits of raw data to determine slope at x_0_ (mean ± standard deviation). Some curves were not able to be fit because they did not come up at all, had no upper baseline, or did not have enough replications (n≥3) to calculate a mean ± standard deviation for the three parameters.

The rate of CSF SOD1 seeding activity at the midpoint (slope), lag phase, or ThT fluorescence amplitude at midpoint (ThT FA_m_) was calculated for ALS CSF and control CSF. ALS CSF SOD1 seeding activity parameters were compared to their ALSFRS-R score (visit 1 and visit 2), their ALSFRS-R slope decline (visit 2 only), and their CSF neurofilament light (visit 1 and visit 2) using linear regression.

A GraphPad Prism outlier test (ROUT, Q = 1%) was used to determine outliers in regression analyses. Sensitivity and specificity was calculated using cut point of 120 hours, and the GraphPad Prism ROC program was used to plot Sensitivity versus 1-Specificity and determine highest likelihood ROC parameters.

## Results

### ALS participants with or without *SOD1* mutations have SOD1 seeding activity in their CSF at initial visit

The SOD1 seed amplification RT-QuIC work flow involves expressing and purifying rSOD1 and mixing with CSF from ALS participants and controls to determine whether SOD1 seeding activity is present (**Fig. 1A**). *SOD1*-ALS constitutes ∼2% of ALS cases and is linked to *SOD1* mutations [33,43,44]. We initially examined premortem CSF for SOD1 seeding activity from five participants clinically diagnosed with *SOD1*-ALS (**Fig. S2A-S2E**) by optimizing our SOD1 RT-QuIC protocol with highly pure rSOD1 WT substrate that has been well characterized [41](**Fig. S1**). Each *SOD1*-ALS participant had a unique *SOD1* mutation, all five *SOD1*-ALS participants were receiving the SOD1 lowering therapy tofersen (Qalsody), and *SOD1*-ALS participants 4 and 5 were additionally receiving riluzole (Rilutek) (**Table 1**).

**Fig 1A.**
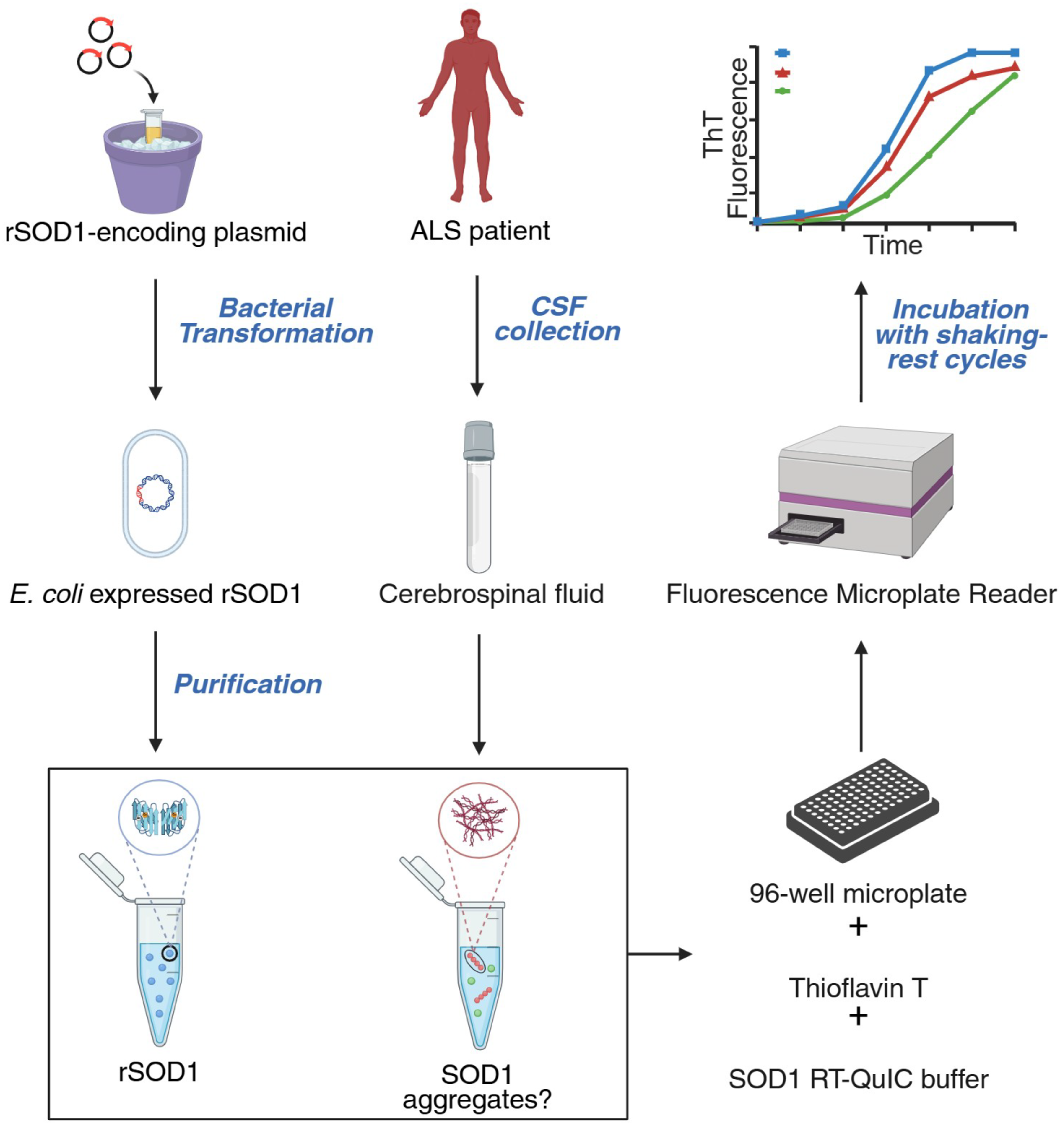
Schematic for detecting SOD1 RT-QuIC seeding activity from ALS participant cerebrospinal fluid.

At the initial CSF collection timepoint (range: 65-168 months from symptom onset), CSF SOD1 seeding activity was observed in three of the five *SOD1*-ALS participants via increased ThT fluorescence intensity but not in two age-matched controls (including CSF from an ALS asymptomatic participant harboring a *C9ORF72* mutation), and a negative non-CSF control containing rSOD1 with ThT and buffer (**Fig. 1B and Figs. S2A-S2E)**. We next examined potential SOD1 seeding activity in premortem CSF from sALS CSF samples (n=13) from participants who were determined by whole exome sequencing to have no mutations in the following genes: Superoxide Dismutase 1 (*SOD1*), Chromosome 9 opening reading frame 72 (*C9ORF72*), Transactivation Response DNA Binding protein of 43 kD (*TARDBP*), Fused in sarcoma (FUS), tau (*MAPT*), Angiogenin (*ANG*), Valosin-containing protein (*VCP*), Vesicle-associated membrane-associated protein (*VAPB*), Progranulin (*GRN*), and Senataxin (*SETX*) (**Table 2**).

**Fig. 1B.**
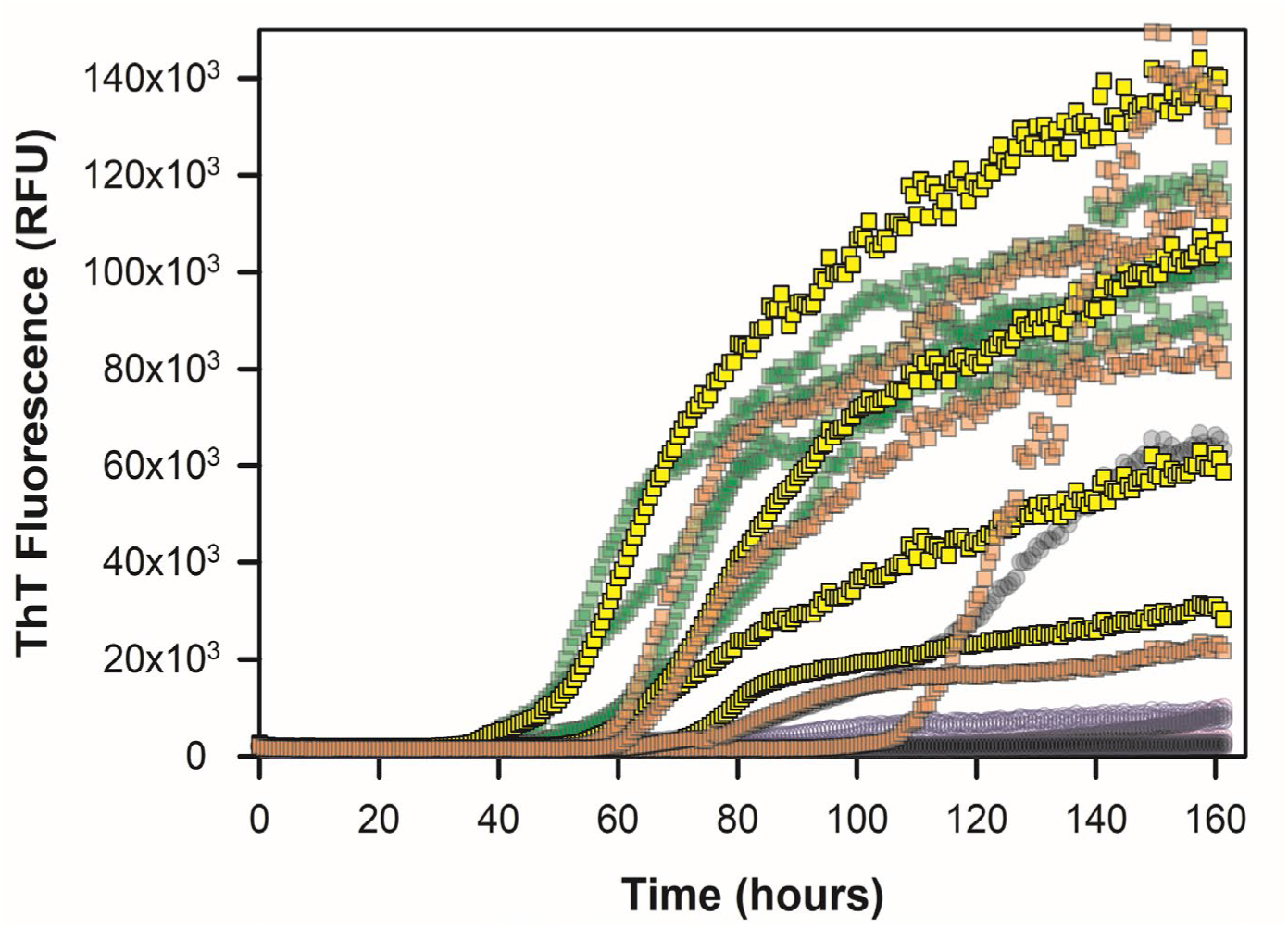
Plot of CSF SOD1 seeding activity from *SOD1*-ALS participants 1 (green squares), 2 (orange squares), and 3 (yellow squares) versus CSF from controls 1 (closed black circles) and 15 (open pink circles) at their initial visit (visit 1). Reaction mix alone (open blue circles) is shown as non-CSF control.

At their initial CSF collection timepoint (range: 13-111 months from symptom onset), several of these ALS participants also displayed SOD1 seeding activity in their CSF, not observed in two controls, or rSOD1 with ThT and buffer only negative control, suggesting WT SOD1 can become aggregation prone in the broader ALS population (**Fig. 1C and Figs. S3A-S3M**).

**Fig. 1C.**
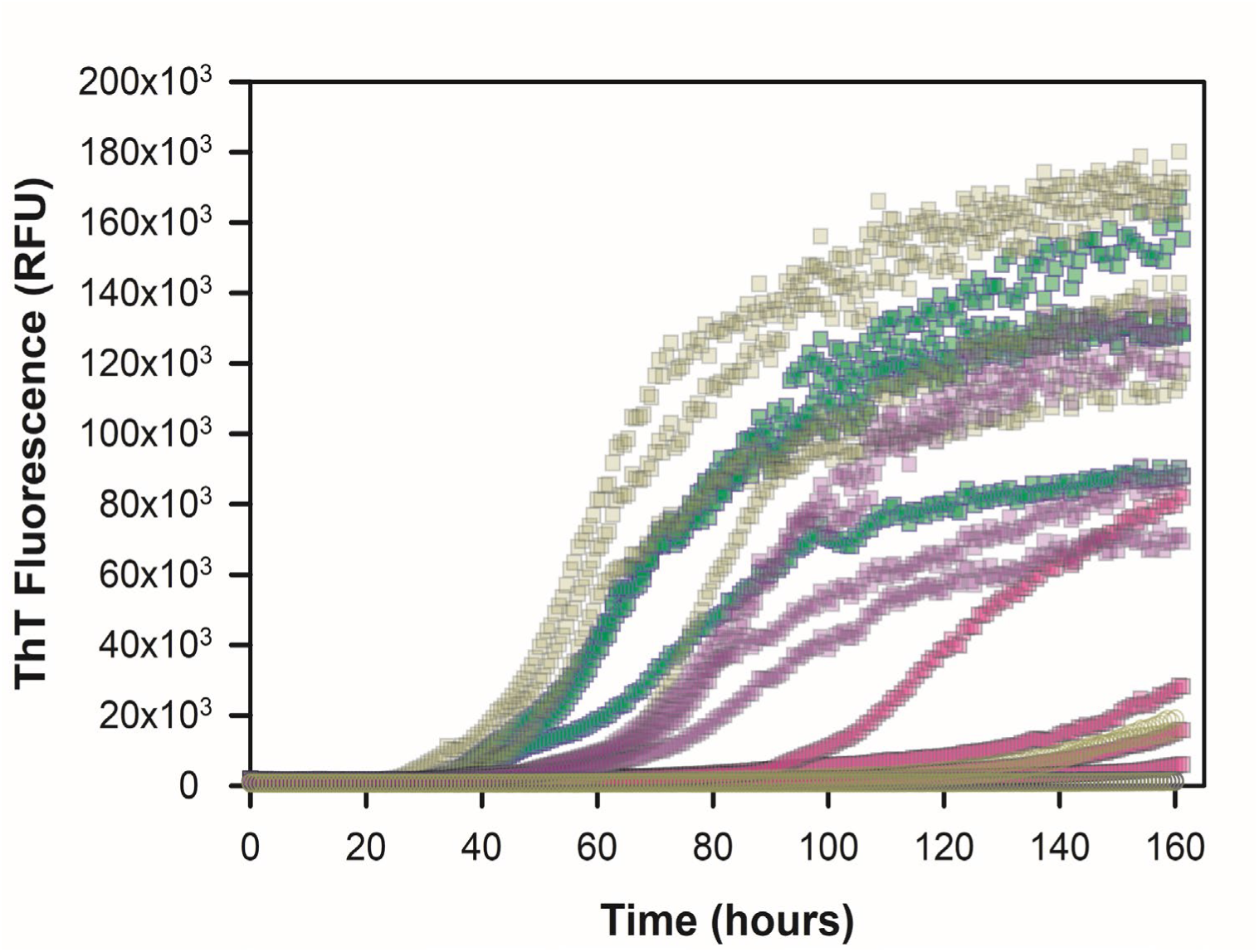
Plot of CSF SOD1 seeding activity from sALS participants 2 (green squares), 3 (dark yellow squares), 6 (purple squares), 10 (pink squares) versus CSF from controls 8 (closed black circles) and 12 (open grey circles) at their initial visit (visit 1). Reaction mix alone (open blue circles) is shown as non-CSF control.

Most healthy controls (**Figs. S4A-S4K, S5**) did not exhibit major ThT fluorescence intensity in their CSF. However, asymptomatic *C9ORF72* mutations carriers (**Figs. S6A-S6C**) showed minimal to moderate increases in ThT fluorescence intensity at subsequent visit without clinical indication of phenoconversion. Moreover, using an antibody to misfolded SOD1 in ALS CSF, we observed incubation of ALS CSF with misfolded SOD1 specific antibody still detected SOD1 seeding activity, but not in the isotype control antibody as it interferes with seeded conversion process (**Figure S7**), suggesting specificity of the assay for misfolded SOD1. An ROC curve was generated to classify control (n=32) and all ALS participants (n=23) at earliest visit using ThT fluorescence and lag phase measured in CSF (threshold 5,000 RFU and 120 hours), producing 80% sensitivity, 92% specificity, and area under the curve equal to 0.95 (**Figure 1D, Table 3**).

**Table 3.** Receiver operator characteristic plot of CSF SOD1 RT-QuIC assay from ALS participants and controls.

| CSF from ALS participants and controls |  |  |  |  |  |  |  |
| --- | --- | --- | --- | --- | --- | --- | --- |
| Area under ROC curve |  |  | sensitivity | 95% CI | specificity | 95% CI | likelihood ratio |
| Area | 0.9516 | >0.125 | 100.0% | 93.0-100% | 58.82% | 45.1-71.2% | 2.429 |
| std error | 0.01803 | >0.375 | 92.16% | 81.5-96.9% | 80.39% | 67.5-88.9% | 4.70 |
| **95% CI** | **0.91-0.98** | **>0.625** | **80.39%** | **67.5-88.9%** | **92.16%** | **81.5-96.9%** | **10.25** |
| p value | <0.0001 | >0.875 | 58.82% | 45.1-71.25% | 100.0% | 93.0-100% | - |

**Fig. 1D.**
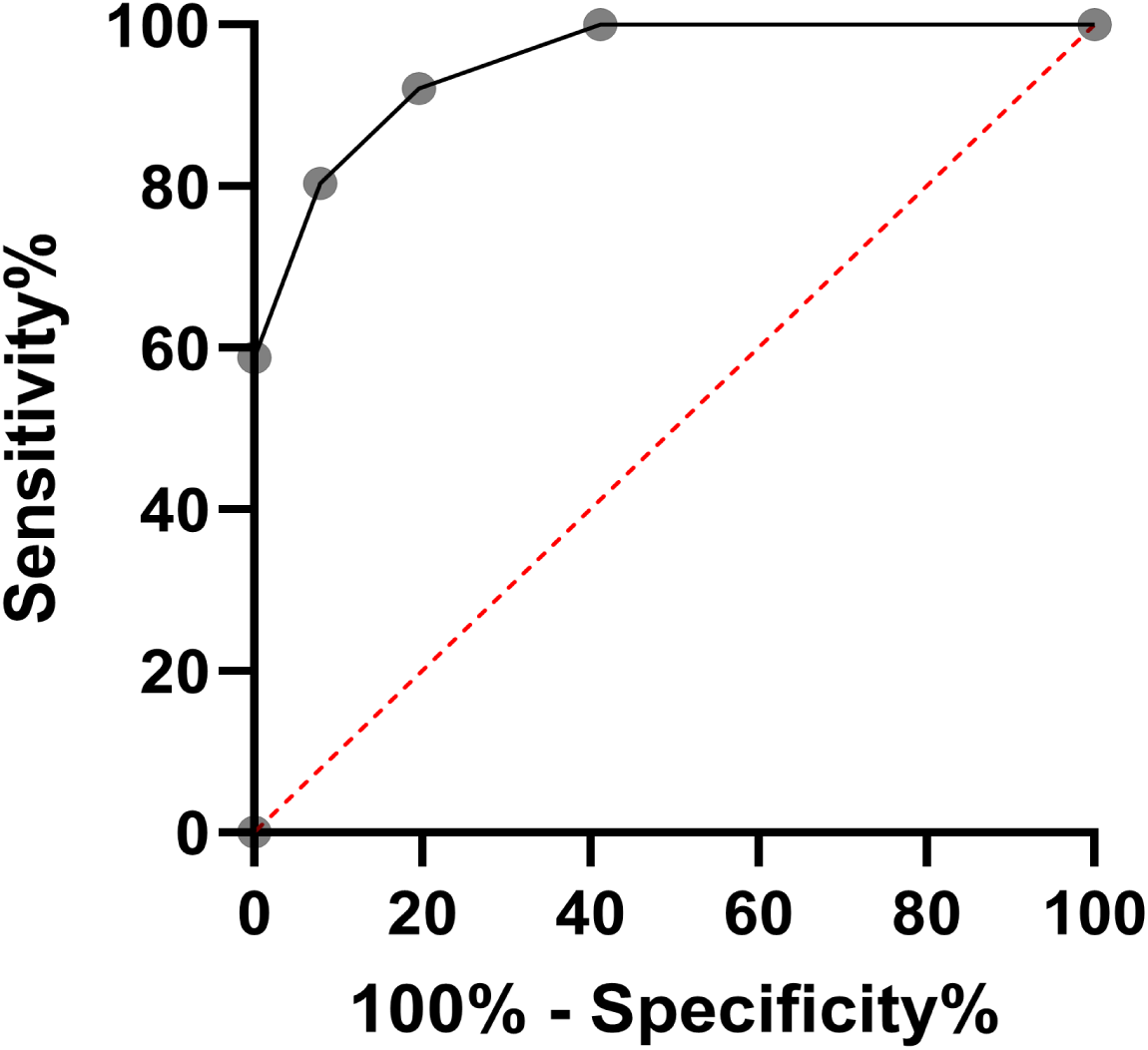
Receiver operator characteristic (ROC) plot for CSF SOD1 seeding activity from 23 ALS participants and 32 controls using threshold of 5,000 RFU and 120 hours at earliest visit.

We also measured CSF SOD1 RT-QuIC seeding activity from participants with other neurodegenerative or neurological diseases, such as spinal muscular atrophy type 3, another motor neuron disease and clinical ALS mimic (n=4, mean RFU 344 ± 1,228, **Fig. S8A**), Parkinson’s disease (n=1, mean RFU 24,875 ± 8,062, **Fig. S8B**), Alzheimer’s disease (n=1, mean RFU 24,071 ± 6,745, **Fig. S8B**), Frontotemporal Dementia (n=4, mean RFU 12,377 ± 15,733, **Fig. S9A**), anxiety (n=2, mean RFU 9,955 ± 8,793, **Fig. S9B**), human immunodeficiency virus (n=3, mean RFU 4,986 ± 5,389, **Fig. S9C**), and mild cognitive impairment (n=1, mean RFU 220 ± 169, **Fig. S8A**) (**Table 1**), and found that the ThT fluorescence amplitudes did not reach levels seen in CSF samples from *SOD1*-ALS (n=5, mean RFU 64,847.1, **Fig. 2C**) and sALS (n=13, mean RFU: 77,041.4, **Fig. 2D**) participants. However, there were some exceptions seen with CSF from AD, PD, and FTD, though ThT fluorescence amplitudes did not exceed 40,000 RFU (**Fig. S8B-S9A**). The majority of CSF from disease controls however, were found to be ThT negative, based on our threshold conditions of 5,000 RFU and 120 hours. Overall, our results suggest the misfolding of SOD1 (wildtype or mutant) occurs in ALS, whether linked to *SOD1* mutations or not. Moreover, in a subset of ALS, SOD1 seeding was observed to occur < 24 months from symptom onset, suggesting that this can be observed early during the disease course (**Figs. S3B, S3H-J, S3L-M**). We next sought to understand how the seeding activity of mutant or wildtype SOD1 could change during a participant’s disease progression (**Table 4**).

**Table 4.** ALS participant clinical information.

| Participant | ALSFRS score visit 1 | ALSFRS score visit 2 | ALSFRS-R slope (decline/month) |
| --- | --- | --- | --- |
| *SOD1*-ALS 1 | 38 | 38 | 0 |
| *SOD1*-ALS 2 | 43 | 44 | 0.0640 |
| *SOD1*-ALS 3 | 33 | 35 | 0.103 |
| *SOD1*-ALS 4 | 38 | 33 | -0.493 |
| *SOD1*-ALS 5 | 34 | 20 | -2.99 |
| sALS 1 | 39 | 35 | -0.328 |
| sALS 2 | 37 | 28 | -0.675 |
| sALS 3 | 21 | 12 | -0.512 |
| sALS 4 | 34 | 32 | -0.508 |
| sALS 5 | 32 | 27 | -1.66 |
| sALS 6 | 28 | 32 | 0.237 |
| sALS 7 | 41 | 37 | -0.241 |
| sALS 8 | 36 | 35 | -0.142 |
| sALS 9 | 36 | 31 | -0.701 |
| sALS 10 | 45 | 39 | -0.814 |
| sALS 11 | 29 | 25 | -1.58 |
| sALS 12 | 44 | 40 | -0.731 |
| sALS 13 | 38 | 30 | -2.11 |
| sALS 14 | 43 | - | -0.236 |
| sALS 15 | 46 | - | 0.490 |
| sALS 16 | 45 | - | -3.44 |
| sALS 17 | 32 | - | -1.24 |
| sALS 18 | - | - | - |

We obtained premortem CSF from the same ALS participants shown in **Figs. 1B–C** from later in their disease course (range: 3-24 months after their initial CSF collection timepoint) and tested aliquots in our CSF SOD1 RT-QuIC assay. Comparing their subsequent visit to their initial visit with *SOD1*-ALS, we observed minor changes in their CSF SOD1 seeding activity kinetics based on the mean (**Fig. 2A and Figs. S2A-S2E**). For the sporadic ALS participants, we observed noticeable changes when comparing their subsequent visit to their initial clinical visit based on the mean (**Fig. 2B and Figs. S3A-S3M**).

**Fig. 2A.**
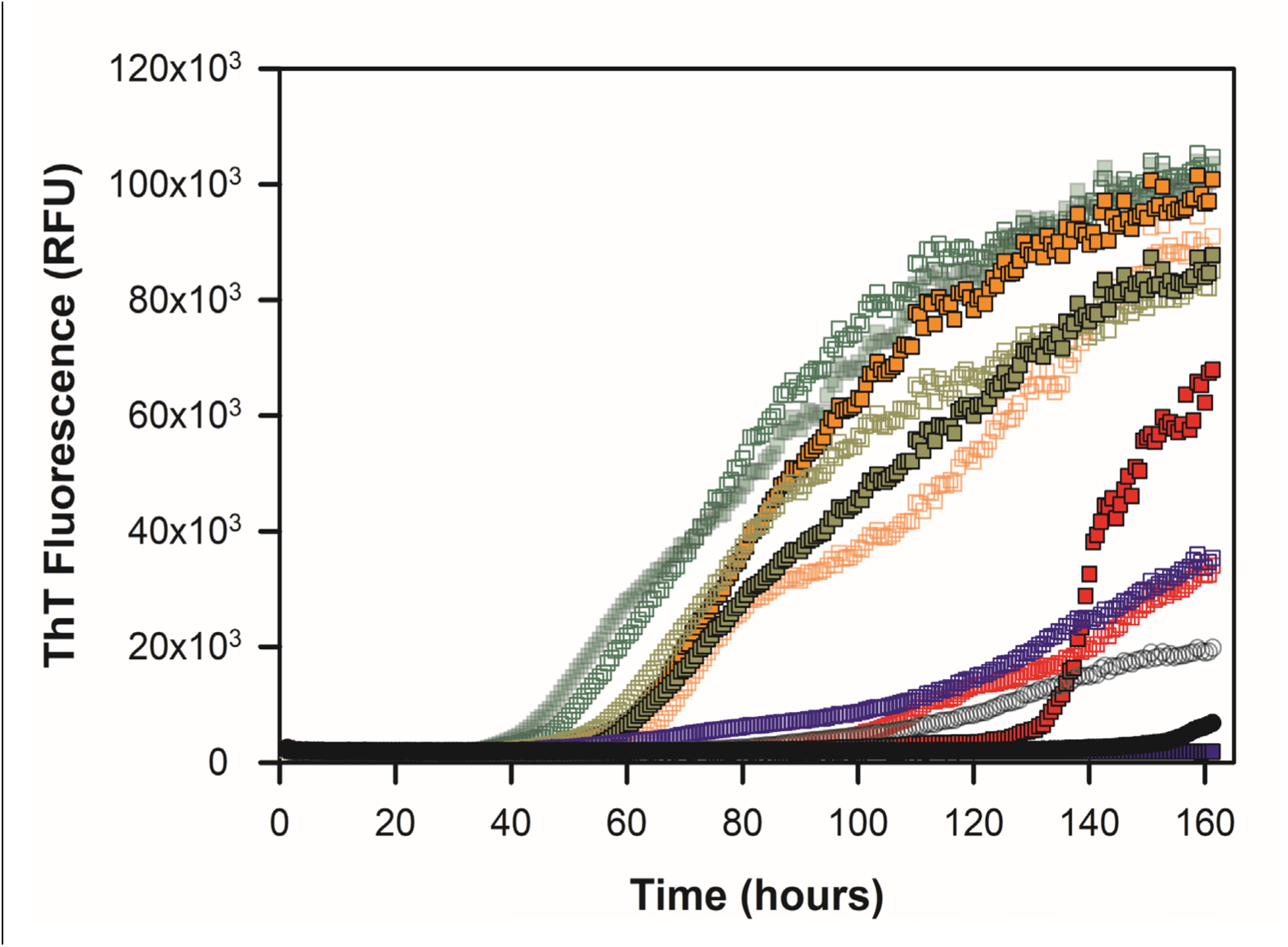
Mean SOD1 RT-QuIC plot of four cerebrospinal fluid replicates from *SOD1*-ALS participants 1 (green squares), 2 (orange squares), 3 (dark yellow squares), 4 (red squares), and 5 (purple squares) versus control 1 (black) at initial visit (visit 1, open symbols) and subsequent visit (visit 2, closed symbols).

**Fig. 2B.**
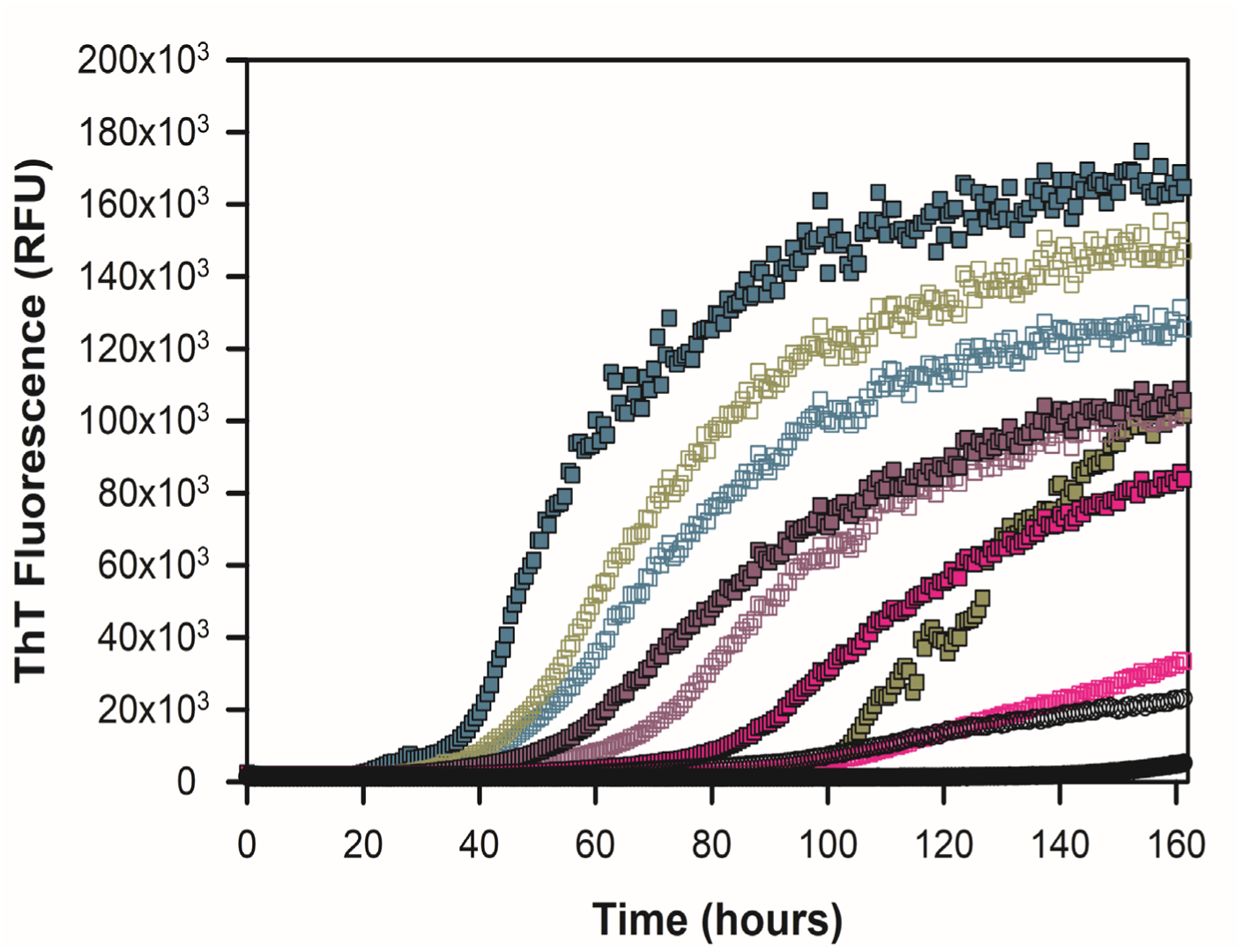
SOD1 RT-QuIC mean plot of four cerebrospinal fluid replicates from sporadic ALS participants 2 (dark cyan squares), 3 (dark yellow squares), 6 (light purple squares), and 10 (pink squares) versus control 11 (black circles) at their first visit (visit 1, open symbols) and subsequent visit (visit 2, closed symbols).

We plotted ThT fluorescence intensity amplitude (final – initial) for all *SOD1*-ALS CSF, sporadic ALS CSF, and control CSF samples from same plates with quadruplicate replication. CSF from *SOD1*-ALS participants (n=5) displayed increased ThT fluorescence amplitude relative to controls (n=6) (*SOD1*-ALS mean: 64,847.1 ± 31,355 RFU, control mean: 16,329.3 ± 15,859 RFU, p-value = 0.009) (**Figure 2C**). ThT fluorescence amplitude was also elevated in sporadic ALS CSF (n=13) compared to controls (n=5) (sALS mean: 77,041.4 ± 41,732 RFU, control mean: 6,711.9 ± 8,957 RFU, p-value = 0.002) (**Figure 2D**). We also plotted ThT fluorescence amplitude of their subsequent visit CSF SOD1 seeding data for *SOD1*-ALS participants (n=5) and controls (n=6) (*SOD1*-ALS mean: 67,895 ± 39,955 RFU, control mean: 45,203 ± 39,704 RFU, p-value = 0.373) (**Figure 2E**) and sporadic ALS participants (n=13) relative to controls (n=5) (sALS median: 99,252 RFU, control median: 18,953 RFU, p-value = 0.004) (**Figure 2F**), respectively. Thus, the replicate mean of sALS participants for their initial and subsequent visits changes more significantly than *SOD1*-ALS participants at both visits.

**Fig. 2C.**
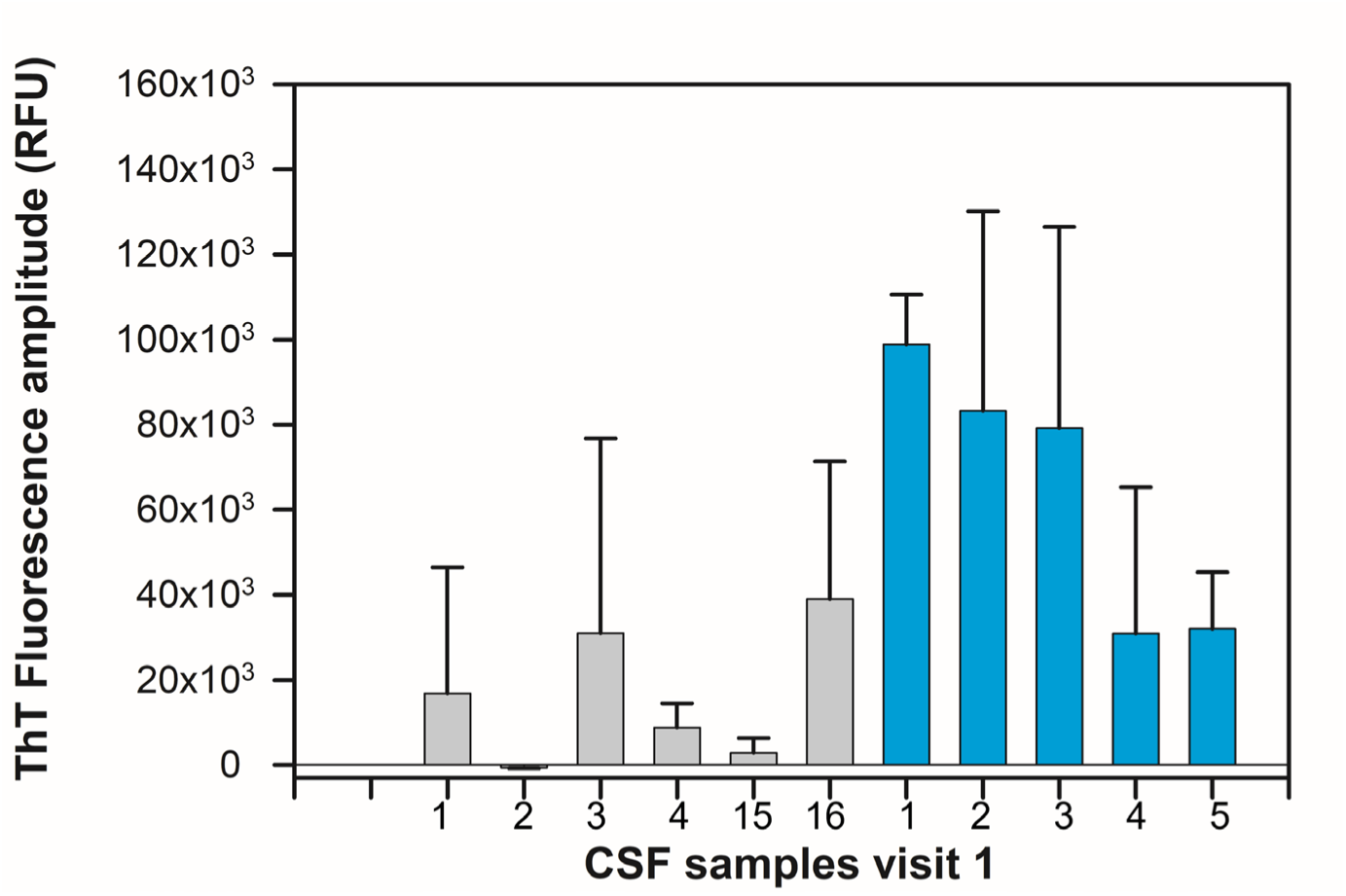
ThT Fluorescence amplitude from *SOD1*-ALS participants (cyan) and controls (grey) at initial visit (visit 1)(p<0.05)

**Fig. 2D.**
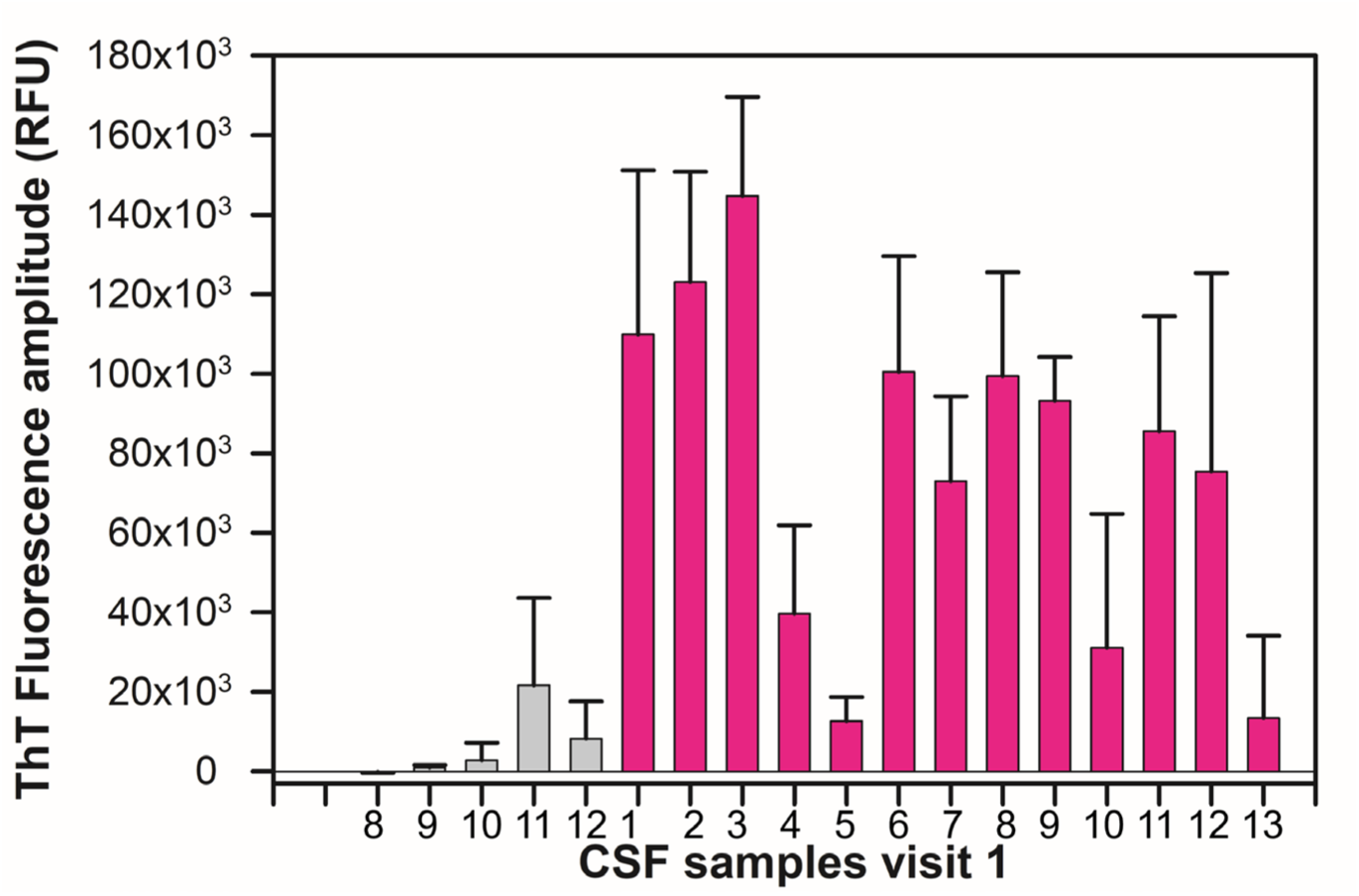
ThT Fluorescence amplitude from sALS participants (pink) and controls (grey) at initial visit (visit 1)(p<0.05).

**Fig. 2E.**
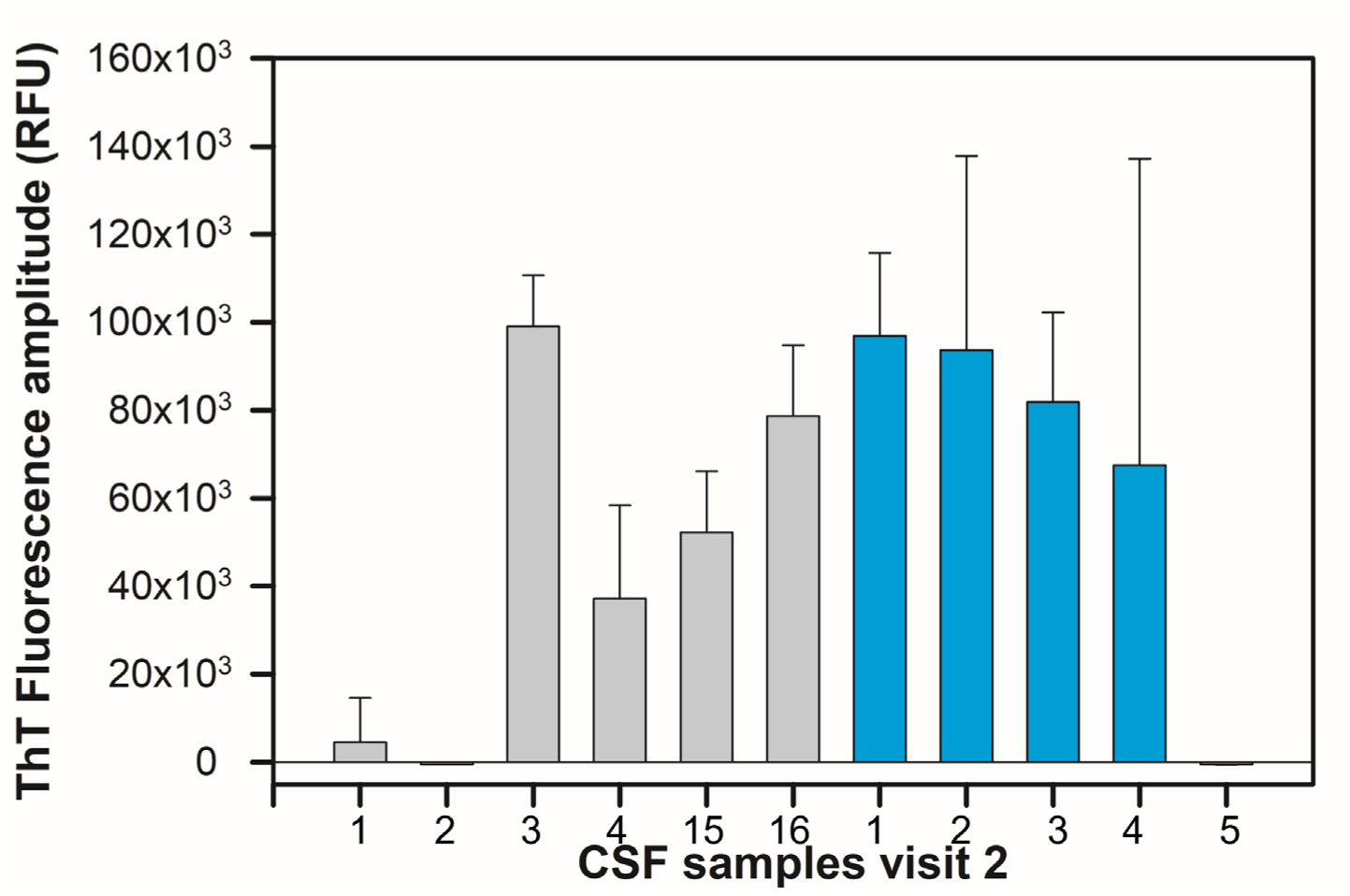
ThT Fluorescence amplitude from *SOD1*-ALS participants (cyan) and controls (grey) at subsequent visit (visit 2).

**Fig. 2F.**
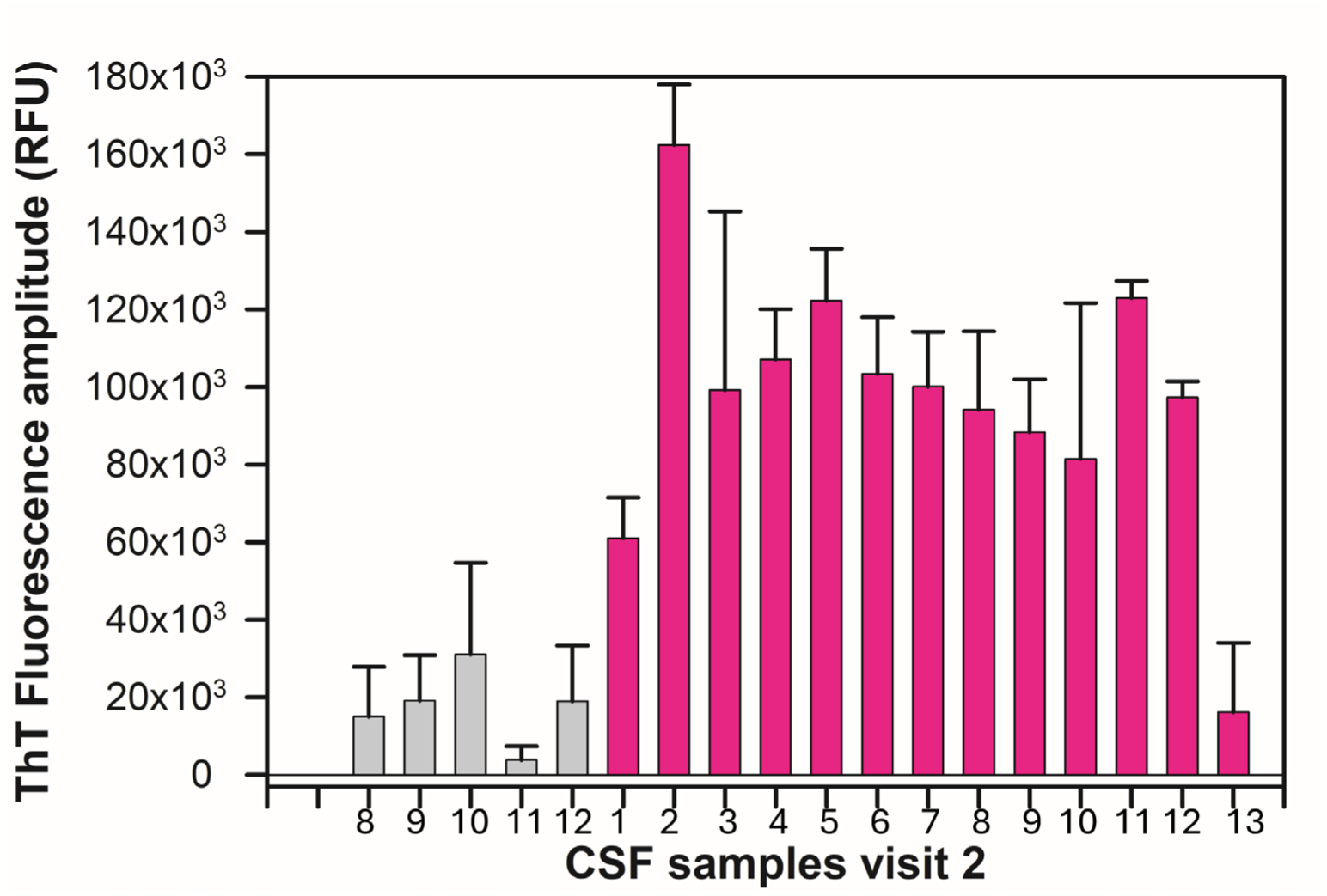
ThT Fluorescence amplitude from sALS participants (pink) and controls (grey) at subsequent visit (visit 2)(p<0.05).

To estimate ALS participant’s CSF SOD1 RT-QuIC parameters, we fit the raw data to **Eq 1** (**Fig S9D**, see Methods) to estimate their ThT fluorescence intensity amplitude at the midpoint (ThT FA_m_), time to detection or lag phase, and the fibril time constant τ, as previously described [41]. We also took the first derivative of **Eq 1** to derive **Eqs 2 and 3** to estimate the CSF SOD1 seeding activity slope at the midpoint. From the fits, we used ThT fluorescence amplitude and the fibril time constant τ to estimate the slope at the midpoint (A/4τ), to produce the rate of CSF SOD1 seeding activity (units in RFU per hour)(**Table 5**).

**Table 5.** CSF SOD1 RT-QuIC parameters (mean ± standard deviation) from ALS participants. Dash symbol indicates their CSF data was unable to be fit to Eq 1.

| ALS participants | Lag phase (hours) |  | SOD1 RT-QuIC slope (RFU/hr) |  | ThT Fluorescence amplitude midpoint (RFU) |  |
| --- | --- | --- | --- | --- | --- | --- |
|  | *Visit 1* | *Visit 2* | *Visit 1* | *Visit 2* | *Visit 1* | *Visit 2* |
| *SOD1* ALS 1 | 50 ± 10 | 44 ± 2 | 1,814 ± 394 | 1,367 ± 390 | 48,565 ± 5,138 | 51,693 ± 9,706 |
| *SOD1* ALS 2 | 71 ± 29 | 58 ± 8 | 2,033 ± 1,678 | 1,723 ± 850 | 44,675 ± 26,731 | 47,518 ± 21,923 |
| *SOD1* ALS 3 | 54 ± 6 | 65 ± 15 | 1,303 ± 826 | 1,299 ± 336 | 39,444 ± 22,232 | 43,320 ± 6,362 |
| *SOD1* ALS 4 | 109 ± 26 | - | 1,304 ± 183 | - | 28,706 ± 20,349 | - |
| *SOD1* ALS 5 | 65 ± 13 | - | 386 ± 207 | - | 19,128 ± 10,193 | - |
| sALS 1 | 58 ± 8 | - | 1,583 ± 477 | - | 58,242 ± 21,438 | - |
| sALS 2 | 44 ± 2 | 32 ± 5 | 2,035 ± 434 | 3,166 ± 322 | 62,137 ± 13,380 | 80,287 ± 6,413 |
| sALS 3 | 44 ± 12 | 104 ± 9 | 2,788 ± 601 | 3,518 ± 2,126 | 71,411 ± 12,739 | 66,404 ± 18,745 |
| sALS 4 | - | 34 ± 5 | - | 1,392 ± 352 | - | 56,059 ± 7,113 |
| sALS 5 | - | 44 ± 2 | - | 1,687 ± 251 | - | 62,610 ± 6,100 |
| sALS 6 | 61 ± 5 | 49 ± 3 | 1,655 ± 637 | 1,498 ± 224 | 51,880 ± 14,364 | 52,914 ± 6,577 |
| sALS 7 | 82 ± 4 | 74 ± 18 | 1,297 ± 373 | 1,618 ± 154 | 39,427 ± 9,624 | 50,666 ± 4,957 |
| sALS 8 | 42 ± 4 | 59 ± 7 | 1,304 ± 207 | 1,369 ± 176 | 49,575 ± 12,584 | 47,738 ± 8,917 |
| sALS 9 | 42 ± 7 | 63 ± 2 | 1,346 ± 200 | 1,309 ± 148 | 45,804 ± 4,511 | 44,189 ± 7,316 |
| sALS 10 | - | 77 ± 5 | - | 1,344 ± 678 | - | 43,849 ± 20,728 |
| sALS 11 | 66 ± 14 | 42 ± 3 | 1,446 ± 361 | 2,092 ± 227 | 44,780 ± 14,755 | 62,129 ± 2,874 |
| sALS 12 | 71 ± 9 | 70 ± 3 | 1,482 ± 196 | 1,515 ± 120 | 57,151 ± 6,378 | 51,850 ± 3,379 |
| sALS 13 | - | - | - | - | - | - |
| sALS 14 | - | - | - | - | - | - |
| sALS 15 | 99 ± 1 | - | 410 ± 141 | - | 23,426 ± 4,758 | - |
| sALS 16 | 85 ± 2 | - | 445 ± 31 | - | 27,064 ± 1,379 | - |
| sALS 17 | - | - | - | - | - | - |
| sALS 18 | - | - | - | - | - | - |

| Controls | Lag phase (hours) |  | SOD1 RT-QuIC Slope (RFU/hr) |  | ThT Fluorescence Amplitude at Midpoint (RFU) |  |
| --- | --- | --- | --- | --- | --- | --- |
|  | *Visit 1* | *Visit 2* | *Visit 1* | *Visit 2* | *Visit 1* | *Visit 2* |
| Control 1 | - | - | - | - | - | - |
| Control 2 | - | - | - | - | - | - |
| Control 3 | - | 57 ± 12 | - | 1,472 ± 156 | - | 51,100 ± 6,008 |
| Control 4 | - | - | - | - | - | - |
| Control 5 | - | - | - | - | - | - |
| Control 6 | - | - | - | - | - | - |
| Control 7 | - | - | - | - | - | - |
| Control 8 | - | - | - | - | - | - |
| Control 9 | - | 65 ± 24 | - | 420 ± 294 | - | 9,890 ± 5,531 |
| Control 10 | - | 62 ± 19 | - | 763 ± 269 | - | 20,200 ± 8,437 |
| Control 11 | - | - | - | - | - | - |
| Control 12 | - | - | - | - | - | - |
| Control 13 | - | - | - | - | - | - |
| Control 14 | - | - | - | - | - | - |
| Control 15 | - | 90 ± 13 | - | 1,060 ± 89.7 | - | 33,900 ± 2,923 |
| Control 16 |  | 83 ± 43 |  | 1,537 ± 156 |  | 43,969 ± 4,186 |
| Control 17 | - | - | - | - | - | - |
| Control 18 | - | - | - | - | - | - |
| Control 19 | - | - | - | - | - | - |
| Control 20 | - | - | - | - | - | - |
| Control 21 | $68 \pm 22$ | - | $355 \pm 80$ | - | $13,200 \pm 3,756$ | - |
| Control 22 | $78 \pm 17$ | - | $454 \pm 110$ | - | $13,500 \pm 500$ | - |
| Control 23 | - | - | - | - | - | - |
| Control 24 | $90 \pm 12$ | - | $254 \pm 154$ | - | $16,036 \pm 7,182$ | - |
| Control 25 | $100 \pm 53$ | - | $182 \pm 128$ | - | $8,506 \pm 5,122$ | - |
| Control 26 | - | - | - | - | - | - |
| Control 27 | - | - | - | - | - | - |
| Control 28 | - | - | - | - | - | - |
| Control 29 | - | - | - | - | - | - |
| Control 30 | - | - | - | - | - | - |
| Control 31 | - | - | - | - | - | - |
| Control 32 | - | - | - | - | - | - |

We first correlated lag phase with the midpoint of ThT fluorescence amplitude and found good correlation at the initial study visit (R=0.61, p-values <0.0001 (y-intercept), 0.01 (slope)) (**Figure 3A**) and their subsequent visit (R=0.76, p-values <0.0001 (y-intercept), 0.001 (slope)) (**Figure 3B**). To determine whether CSF SOD1 seeding activity corresponded to clinical status during ALS progression, we graphed the rate of CSF

**Fig. 3A.**
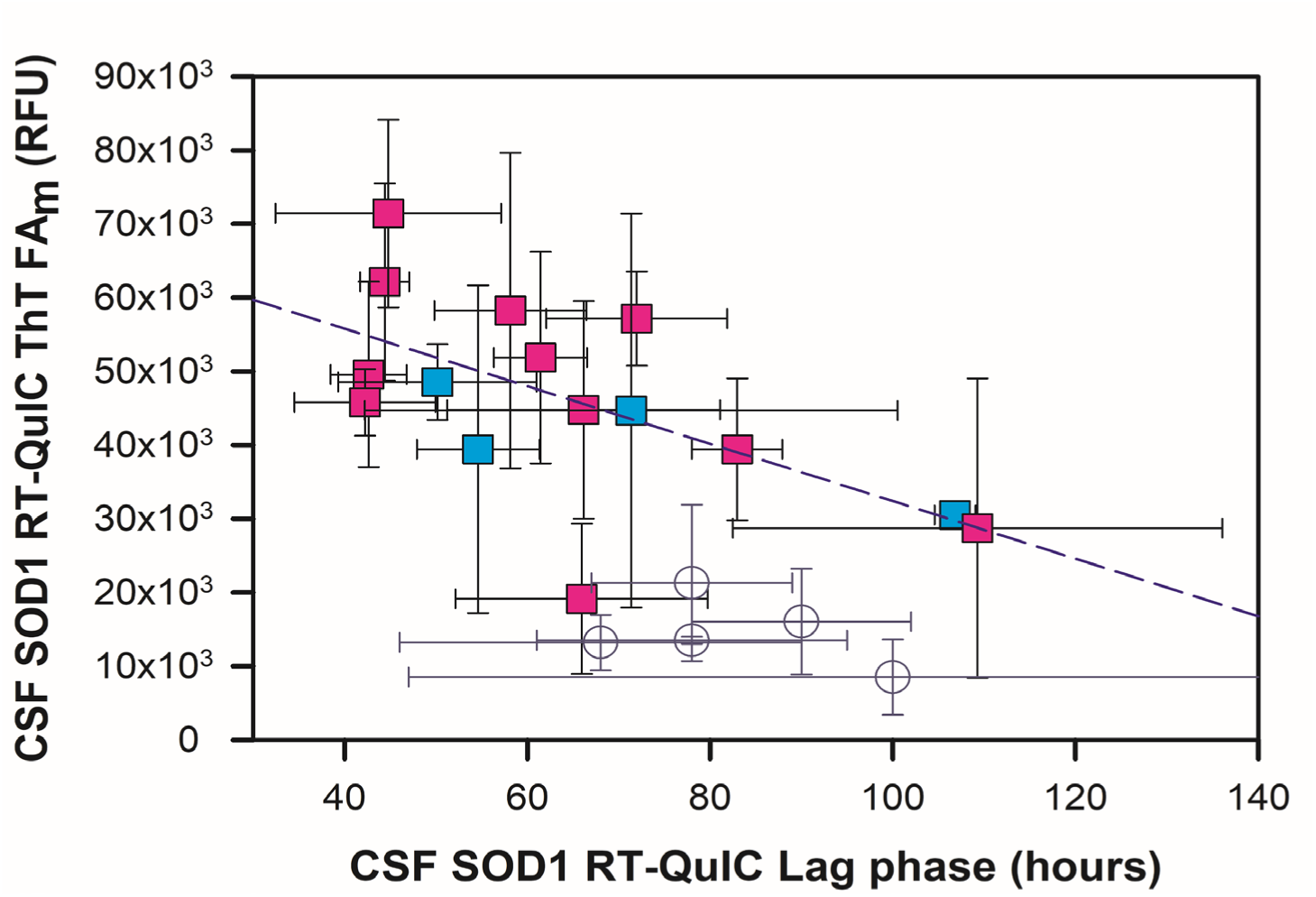
ThT fluorescence intensity amplitude at the midpoint (RFU) versus lag phase (hours) for ALS participant CSF at initial visit (R=0.61, p-values 0.0001 (y-intercept), 0.01 (slope). *SOD1*-ALS in cyan squares, sporadic ALS in pink squares). Controls that gave ThT fluorescence shown in open circles.

**Fig. 3B.**
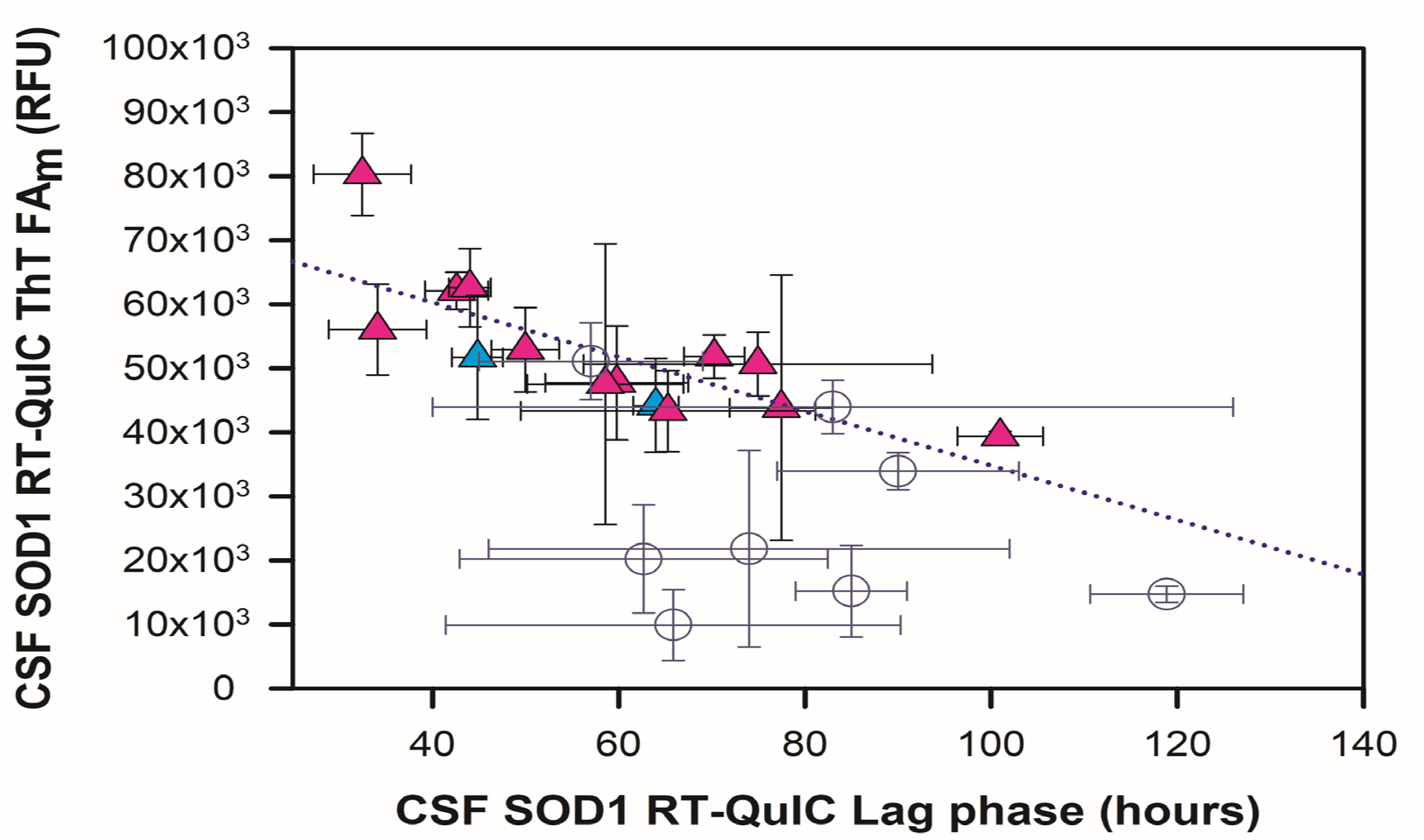
ThT fluorescence intensity amplitude at the midpoint (RFU) versus lag phase (hours) for ALS participant CSF at subsequent visit (R=0.76, p-values 0.0001 (y-intercept) 0.001 (slope). *SOD1*-ALS in cyan triangles, sporadic ALS in pink triangles. Controls that gave ThT fluorescence shown in open circles.

SOD1 seeding activity versus the participant’s ALS Functional Rating Scale (ALSFRS) score and ALSFRS-R slope decline. Sporadic ALS participants 2 and 3 slope values deviated from the trend and were outliers in the ALSFRS-R slope decline analysis assessed with outlier statistical test (see methods, **Figs. S10A**). Removing those two outliers, we observed a significant correlation of the rate of CSF SOD1 RT-QuIC seeding activity versus the ALSFRS score (**Fig. 4A,** R=0.59, p-values 0.0007 (slope) <0.0001 (y-intercept)) and ALSFRS-R slope decline (**Fig. 4B,** R=0.54, p-values 0.055 (slope) <0.0001 (y-intercept)). Thus, the rate of an ALS participant’s CSF SOD1 seeding activity tends to increase with ALS severity and functional decline. We also examined relationships between CSF SOD1 RT-QuIC ThT fluorescence amplitude at midpoint with ALSFRS score from both visits (R=0.46, p-values 0.012 (slope) 0.0001 (y-intercept)) and ALSFRS-R slope decline (R=0.42, p-values 0.11 (slope) <0.0001 (y-intercept))(**Figs. 4C and 4D and Fig. S10B**). Contrarily, lag phase had poor relationship with ALSFRS score (R=0.05, p-values 0.76 (slope) 0.004 (y-intercept)(**Fig.S11**) and ALSFRS-R slope decline (R=0.18, p-values 0.51 (slope) 0.0001 (y-intercept)). Overall, rate of CSF SOD1 RT-QuIC seeding activity correlated the best with ALS progression.

**Fig. 4A.**
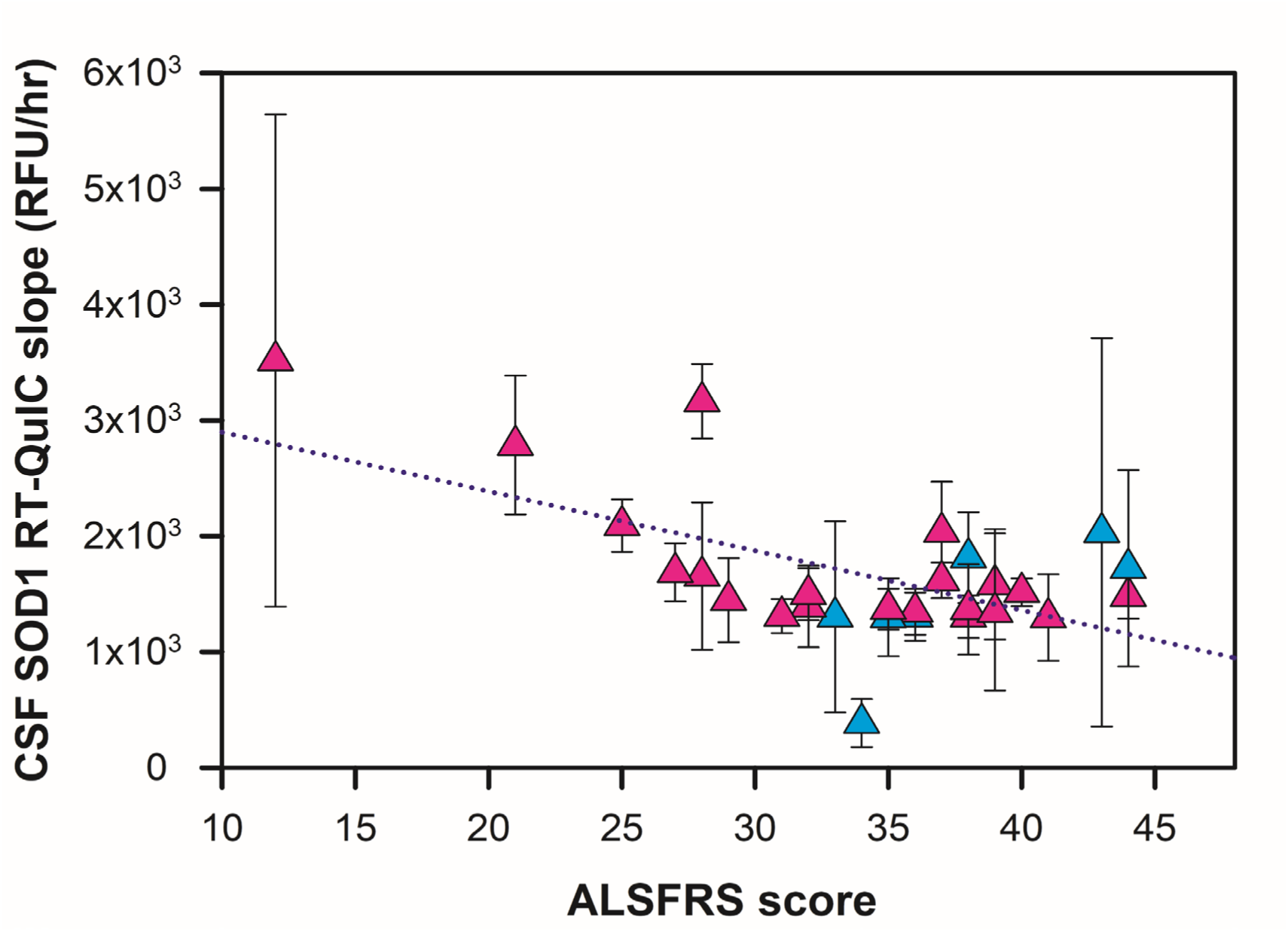
CSF SOD1 RT-QuIC slope (RFU/hr) from ALS participants correlates with their ALSFRS-R score from both visits (R=0.59, p-values 0.0001 (y-intercept), 0.055 (slope)). *SOD1*-ALS cyan triangles, sporadic ALS pink triangles.

**Fig. 4B.**
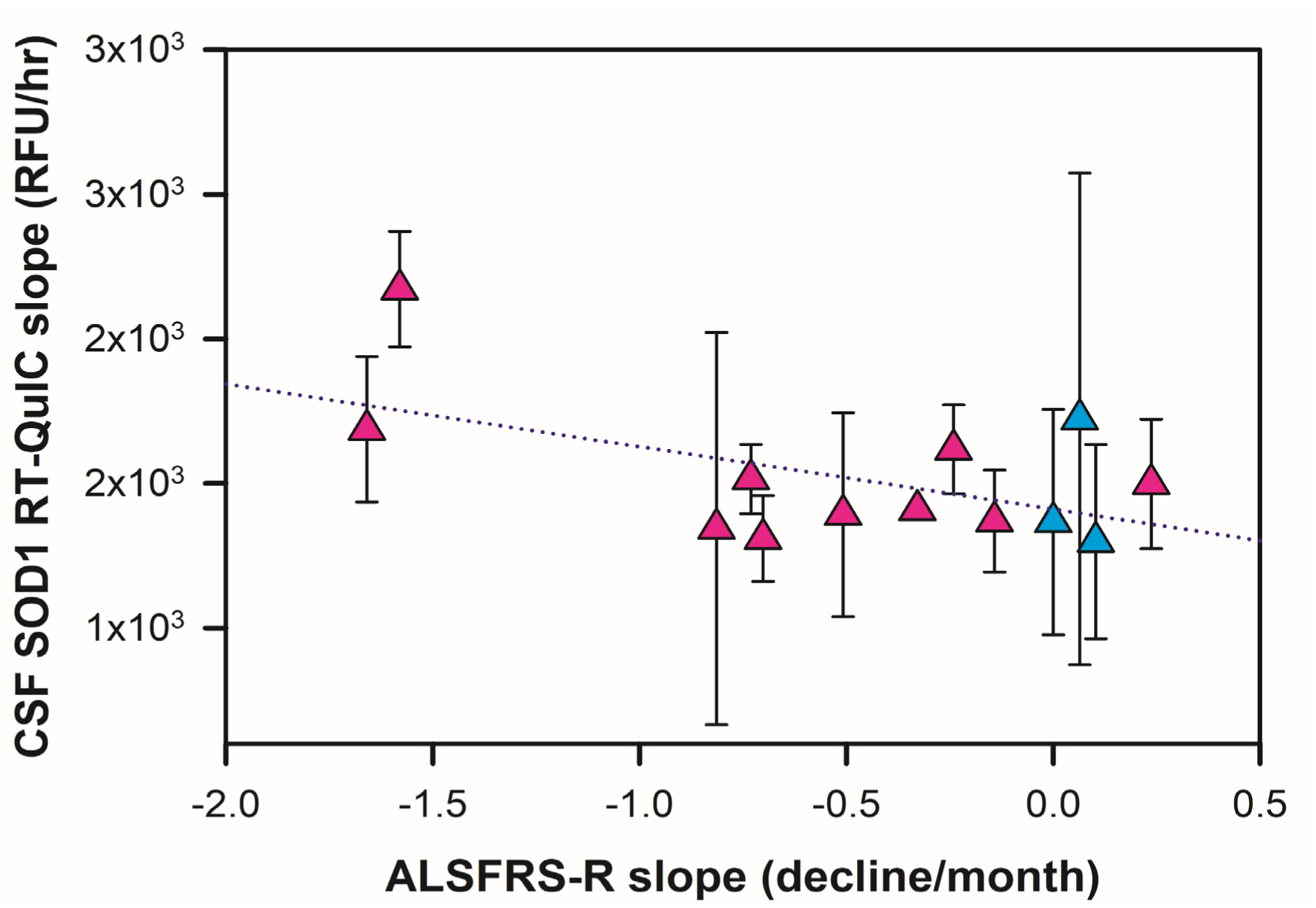
CSF SOD1 RT-QuIC slope (RFU/hr) from ALS participants correlates with their ALS progression (R=0.54, p-values 0.0001 (y-intercept), 0.055 (slope)). *SOD1*-ALS cyan triangles, sporadic ALS pink triangles.

**Fig. 4C.**
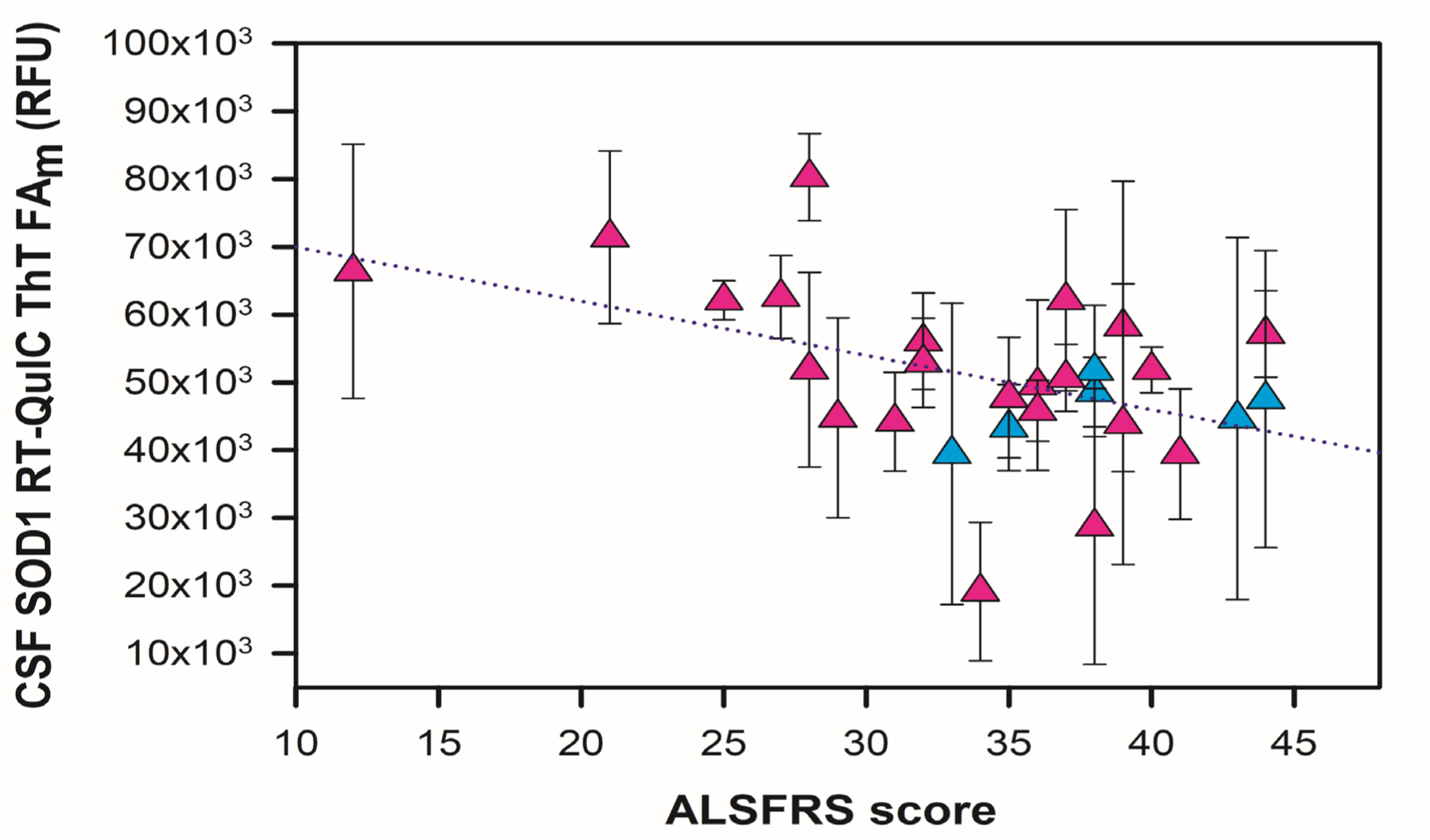
CSF SOD1 RT-QuIC ThT fluorescence amplitude (RFU) from ALS participants versus their ALSFRS score from both visits (R=0.46, p-values 0.0001 (y-intercept), 0.012 (slope)). *SOD1*-ALS cyan triangles and sporadic ALS pink triangles.

**Fig. 4D.**
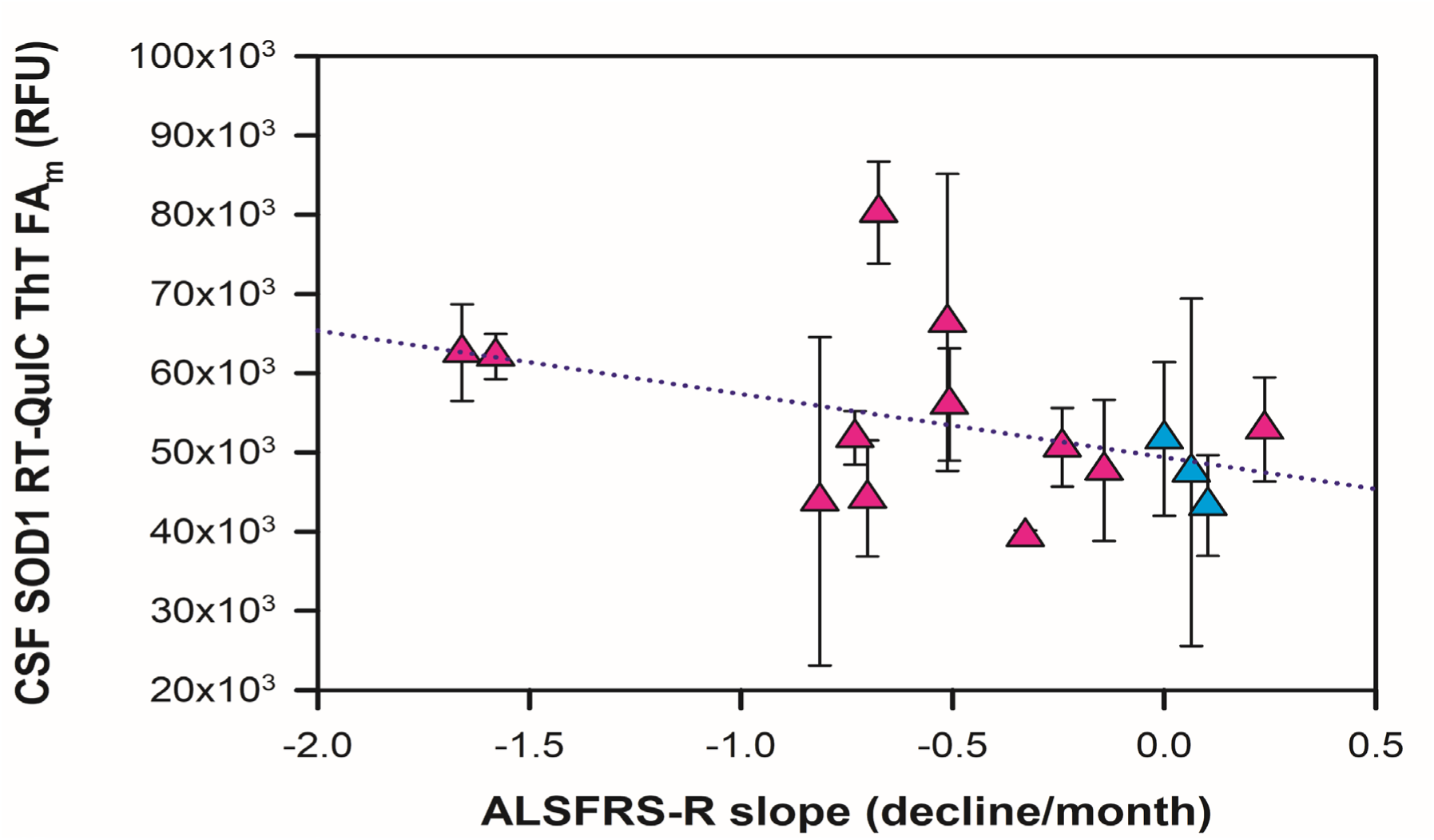
CSF SOD1 RT-QuIC ThT fluorescence amplitude (RFU) from ALS participants versus their ALS progression (R=0.42, p-values 0.0001 (y-intercept), 0.11 (slope)). *SOD1*-ALS cyan triangles and sporadic ALS pink triangles.

To further probe validity of our novel CSF SOD1 RT-QuIC assay, we examined the relationship of CSF SOD1 RT-QuIC parameters with an established ALS CSF biomarker, neurofilament light chain (NfL) [45]. Neurofilament light levels are known to increase early with incipient ALS before peaking soon after diagnosis and has been shown to correlate with ALS disease progression [46]. While the CSF SOD1 RT-QuIC lag phase and slope at midpoint had some trend with CSF Neurofilament light levels (**Fig. S12A-S12B**), CSF SOD1 RT-QuIC ThT fluorescence amplitude at the midpoint at the early and subsequent visits gave the best correlation with CSF neurofilament light chain levels (**Figure 5**, R=0.55, p-values 0.0001 (y-intercept) 0.003 (slope)).

**Fig. 5.**
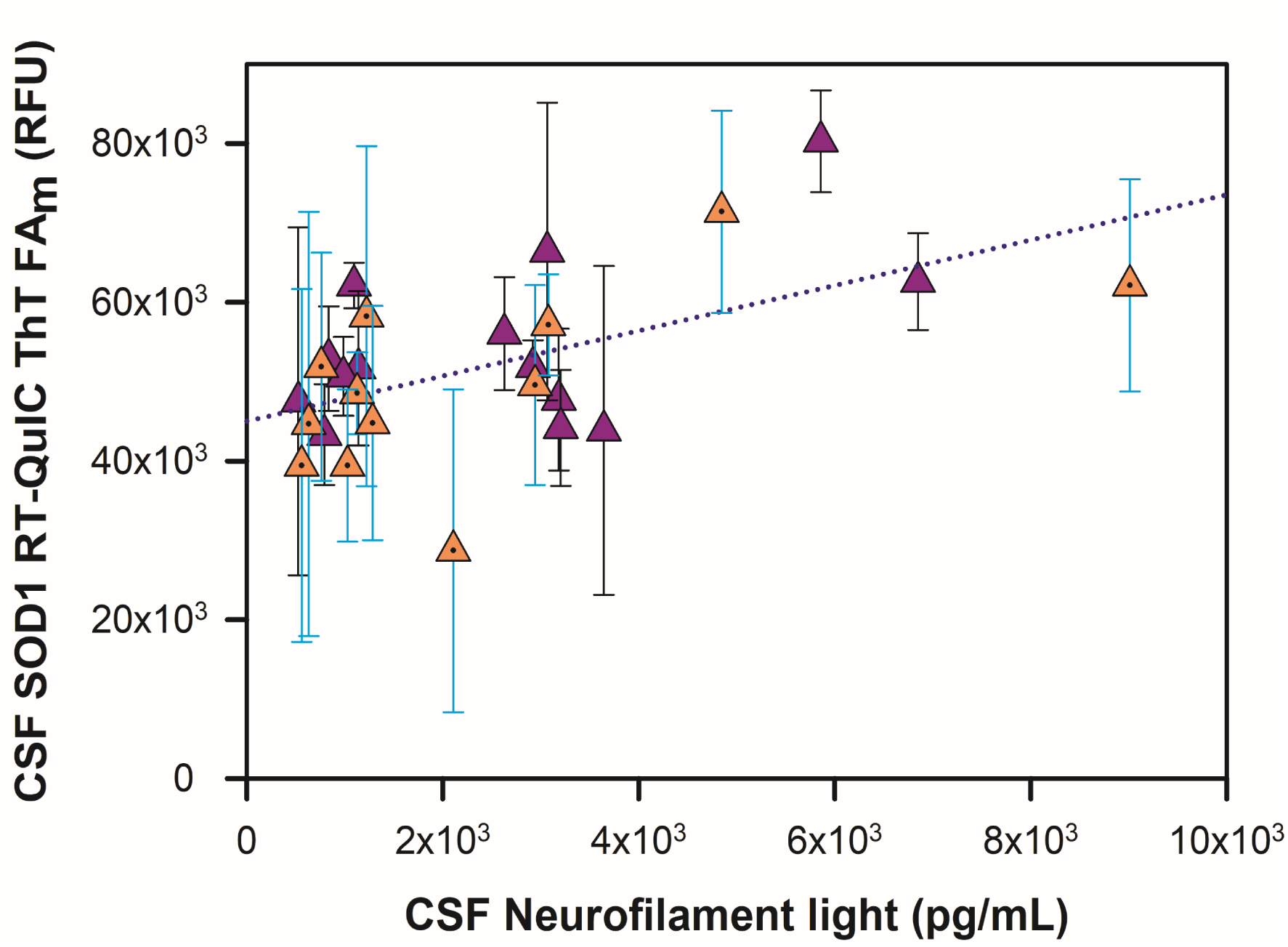
CSF SOD1 ThT fluorescence amplitude (RFU) from ALS participants correlates with their CSF neurofilament light (pg/mL) at visit 1 (orange triangles) and visit 2 (purple triangles). For both visits, R=0.55, p-values 0.0001 (y-intercept), 0.003 (slope)).

In addition to diagnostic and prognostic potential of this antemortem CSF SOD1 RT-QuIC assay, the SOD1 seed amplification assay could also assist with stratification of ALS patients for new clinical trials with SOD1 lowering or other SOD1-targeted therapies, by identifying participants with high SOD1 seeding activity and protein misfolding as a component of their ALS progression. At the initial visit (ALS participant symptom onset to first CSF collection range: 13-168 months), 14/18 ALS CSF samples (3/5 *SOD1*-linked ALS, 11/13 sporadic ALS) displayed CSF SOD1 seeding activity, indicating SOD1 misfolding may not be a universal widespread pathology in ALS. However, integrating CSF data from both initial and subsequent visits, 16/18 ALS participants with longitudinal data examined here who were relatively early in their ALS had CSF seeding competent SOD1. Disease-modifying treatments that have shown stabilization or slowing of disease in *SOD1*-ALS participants [47], might also be beneficial to patients in the broader ALS population who have misfolded SOD1 pathology. Because there are few FDA treatments approved for a majority of ALS patients, the repurposing of tofersen (Qalsody), or the use of other SOD1-targeted therapies for sporadic ALS patients with SOD1 pathology, may be a viable therapeutic strategy in treating a majority of ALS. Nonetheless, it remains unknown whether SOD1 seeding activity reflects a pathogenic process in ALS and/or represents a by-product of cellular oxidative stress and damage seen within neurodegenerative conditions such as ALS.

## Discussion

Many neurodegenerative diseases (NDs) are defined by the abnormal accumulation of misfolded β-sheet-rich protein aggregates that often precede progressive neuron loss and prion-like pathological spread [48,49]. Misfolded proteins in NDs include tau in Alzheimer’s disease and Frontotemporal Dementia (FTD)[50,51], α-synuclein in Parkinson’s disease and Lewy body disease [52], prion in prion diseases [53,54], and TDP-43, FUS, and SOD1 in FTD and ALS [55–59]. Because protein misfolding can occur prior to overt neuronal loss, the emergence of seed amplification assays (SAA) has potential to provide an early window into the neurodegenerative process and help stratify conditions with distinct pathologies *antemortem*. SAAs implementing the RT-QuIC platform have emerged as important methods for studying proteinopathy in neurodegenerative diseases [60], enabling the detection of specific misfolded proteins for potential biomarker development and clinical trial involvement. Several seed amplification RT-QuIC assays are now in use to study proteinopathy, with prion and alpha synuclein RT-QuIC assays currently used the most frequently in clinical settings [61,62]. While other prion-like proteins such as Fused in sarcoma [63], TDP-43 [11,20,57,64,65], and SOD1 [1,19–22,41,43,66,67] have been reported in FTD and ALS pathology, none have been studied during ALS disease progression, and in a diagnostic timeframe when ALS participants with SOD1 pathology could be enrolled in clinical trials (<2 years from symptom onset) as described here.

The timeframe to diagnose ALS is often delayed, particularly if ALS patients have limited access to neurologists such as ALS patients living in rural areas [68]. On average, diagnosing ALS can take 10 – 16 months after their limb or bulbar symptom onset [69], and by the time an ALS patient receives their diagnosis, on average they have 2 – 5 years to live [1]. In many situations with respect to diagnosing ALS, the opportunity for newly diagnosed ALS participants to be involved in new ALS clinical trials is often too late, as the disease has rapidly progressed [70]. Thus, development of new ALS biomarkers is necessary to facilitate timely ALS diagnosis, stratify ALS participants with specific pathology to enroll in new trials, determine their ALS prognosis, enhance our understanding of ALS progression, and examine response to novel disease-modifying therapeutics in new clinical trials with therapies aimed at slowing ALS progression.

In this study, we report that the SOD1 seed amplification RT-QuIC assay used to probe SOD1 misfolding in spinal cord and motor cortices from ALS cases with or without *SOD1* mutations [41], can be used as a potential novel CSF biomarker for ALS participants with or without mutations linked to *SOD1*, ≥90% of all ALS. Many of the antemortem ALS CSF samples analyzed here were collected from sporadic ALS participants that were less than 2 years from their limb or bulbar symptom onset, suggesting this assay has the capability to measure CSF SOD1 seeding activity within typical ALS diagnostic timeframes and possibly aid clinicians in an ALS participant’s involvement in clinical trials with SOD1 therapies. In our initial study, CSF SOD1 seeding activity (wildtype or mutant) was detected in 19 of 23 ALS participants, 3/5 with *SOD1*-ALS and 16/18 with sporadic ALS. Similar CSF ThT fluorescence amplitude enhancements and lag phases were observed from both *SOD1*-linked and non-*SOD1* linked ALS participants, suggesting the mutant and wildtype SOD1 seeds detected in ALS participant antemortem CSF may have similar biochemical properties, such as aggregate concentration, aggregate particle size, shared exposed binding sites in their misfolded SOD1, and the similar capacity of some misfolded mutant SOD1 and wildtype SOD1 conformations in their ability to template our well characterized wildtype human rSOD1 substrate [41].

With respect to SOD1 seeding kinetics in *SOD1*-ALS participant CSF, we observed comparable seeding activity in *SOD1*-ALS participants 1-3 who carry the *SOD1* mutations p.V32A, p.E101K, and p.A90V, which have been reported to have early ALS symptom onset and slow progression [29,71]. However, although *SOD1*-ALS participant 4 harbors a p.L145S mutation that has been described to have similar age of symptom onset and disease duration [72], we did not detect major SOD1 seeding activity in our threshold range for *SOD1*-ALS participant 4. Similarly, *SOD1*-ALS participant 5, who carries an intronic deletion in their *SOD1* gene, did not exhibit major SOD1 seeding activity. Further evaluation of SOD1 seeding activity in a larger cohort of diverse *SOD1* mutations is required to determine the ability of the SOD1 RT-QuIC assay to discern mutation-specific differences in aggregation potential and clinical phenotype.

SOD1 protein consists of 154 amino acids and missense mutations in over 100 residues have been found to cause ALS [27] and have been associated with diverse structural and chemical consequences. Although *SOD1* mutations account for ∼20% of inherited ALS, approximately 1-2% of *SOD1* mutations have been discovered in sporadic ALS cases without evident family history [73–75]. Various factors such as SOD1 stability, dismutase activity, and aggregation potential are not fully predictive of pathogenic potential or clinical phenotype [27,76–78]. Studies have indicated mutant SOD1 co-localizes with 14-3-3 proteins, which can be found in ubiquitinated inclusions in *SOD1*-ALS spinal cords [79], found to form stable complexes [80] and can inhibit 14-3-3 positive cell survival [81]. Thus, there could be other compensatory mechanisms that can influence the progression of *SOD1*-ALS. Future studies examining *SOD1* mutations in these contexts will aid our understanding of *SOD1* mutations linked to ALS pathogenesis, SOD1 seeding activity, and the progression of *SOD1*-ALS.

It is important to note that both *SOD1*-ALS participants 4 and 5 were administered riluzole and tofersen after their ALS diagnosis, while *SOD1*-ALS participants 1-3 were only on tofersen (**Table 1**). Thus, the combination of these therapies among the different *SOD1*-ALS participants studied here confounds the interpretation of CSF SOD1 seeding activity in these individuals. The mechanisms of action of riluzole (Rilutek, glutamate excitotoxicity), edavarone (Radicava, an antioxidant), and tofersen (Qalsody, a SOD1 lowering antisense oligonucleotide), could potentially impact SOD1 seeding activity in the central nervous system. For instance, with respect to the three *SOD1*-ALS participants only on tofersen after their ALS diagnosis, we observed robust CSF SOD1 seeding activity at both clinical visits (**Figures 1 and 2**), with stabilization or reversal in their rate of CSF SOD1 seeding activity at the subsequent clinical visit compared to the first clinical visit (**Figure 5**). Analysis of baseline CSF, prior to therapy initiation, and longitudinal CSF while on SOD1-lowering therapy will be necessary to determine whether SOD1 seeding may serve as a pharmacodynamic response marker. However, only modest CSF SOD1 seeding activity signal was observed in *SOD1*-ALS participants 4 and 5, who were on tofersen and riluzole after their ALS diagnosis (**Figures S2D-S2E**). It is possible that our well characterized human wildtype rSOD1 substrate has less affinity for these misfolded SOD1 conformations in CSF, thus we did not observe efficient propagation, and/or alternatively, there were insufficient or sequestered SOD1 seeds present in their CSF that we could not detect. Determining how CSF SOD1 seeding activity is affected by an ALS participant’s *SOD1* mutation status and subsequently modified by therapeutic interventions will clarify the potential of our CSF SOD1 RT-QuIC assay for several biomarker context of uses across several *SOD1*-ALS participants with varying *SOD1* mutations.

The discovery that antemortem CSF collected from sporadic ALS participants (who have no common risk genetic mutations linked to ALS, **Table 2**) has seeding competent WT SOD1 suggests aberrant WT SOD1 is involved in ALS pathophysiology in the broader population. At least one previous study has reported misfolded WT SOD1 in antemortem CSF via a sandwich ELISA using the monoclonal C4F6 antibody, where the misfolded WT SOD1 in many of these sporadic ALS CSF samples were found to have reduced affinity for copper and zinc [24]. In addition, another study has reported WT SOD1 in ALS postmortem spinal cord is oxidized and misfolded, and shares a common binding site with mutant SOD1 protein, also detected with C4F6 antibody [82]. The human WT rSOD1 substrate used in these ALS CSF studies is in the apo state, contains an intramolecular disulfide bond, and is structurally disordered, to enable detection of SOD1 seeding activity in ALS tissues [41]. Incubation of sporadic ALS CSF with C4F6 antibody increased SOD1 seeding activity via slope (**Figure S7**). Thus, there may be specific post-translational events and modifications in WT SOD1 that occur during sporadic ALS pathogenesis, such as oxidative modifications and the loss of copper and zinc, that could alter the structure of WT SOD1 to a disease associated conformation with seeding activity during ALS pathophysiology. Pathological peroxide concentrations can modify WT SOD1 at cysteine 111, in both recombinant SOD1 and SH-SY5Y cell culture showing fibrillization, and WT SOD1 oxidation has been reported to inhibit kinesin based fast axonal transport [40,82]. Moreover, immune system studies have suggested SOD1 responds to inflammatory stimuli and plays a role in regulating oxidative stress [83–85]. The tryptophan 32 residue is suspected to participate in the WT SOD1 aggregation process [86,87]. Mitochondrial respiration and signaling enzymes produce superoxide, a substrate for SOD1, and disturbances in mitochondrial membrane potential and redox transitions can impact ROS homeostasis, especially after insults such as toxin exposure and ischemia [88,89]. Recent evidence suggests certain environmental factors are associated with elevated ALS risk and shorter ALS survival [70,90–92], and increases in oxidized glutathione have been reported to be elevated in ALS CSF [93]. Thus, aberrant oxidative stress with SOD1 dysfunction could provide an environment where WT SOD1 structure becomes prion-like. Stress at the endoplasmic reticulum in mice has indicated WT SOD1 can aggregate, correlating with astrocyte activation [94]. Recent ALS postmortem tissue studies have reported structurally disordered WT SOD1 conformers being mis-localized in spinal cord motor neurons from sporadic ALS cases [19]. These misfolded forms of SOD1 co-deposit with sequestosome 1 and phosphorylated TDP-43 [20], two key proteins that are pathological hallmarks to motor neuron degeneration. We observed CSF SOD1 RT-QuIC seeding activity in a majority of sporadic ALS participants (**Figures S3A-S3M**), and that ALS CSF SOD1 seeding activity increases via slope and ThT FA_m_ which correlates with the ALSFRS score and ALSFRS-R slope decline (**Figs. 4A-4D**), and CSF neurofilament light (**Fig. 5**)(**Table 6**).

**Table 6.**
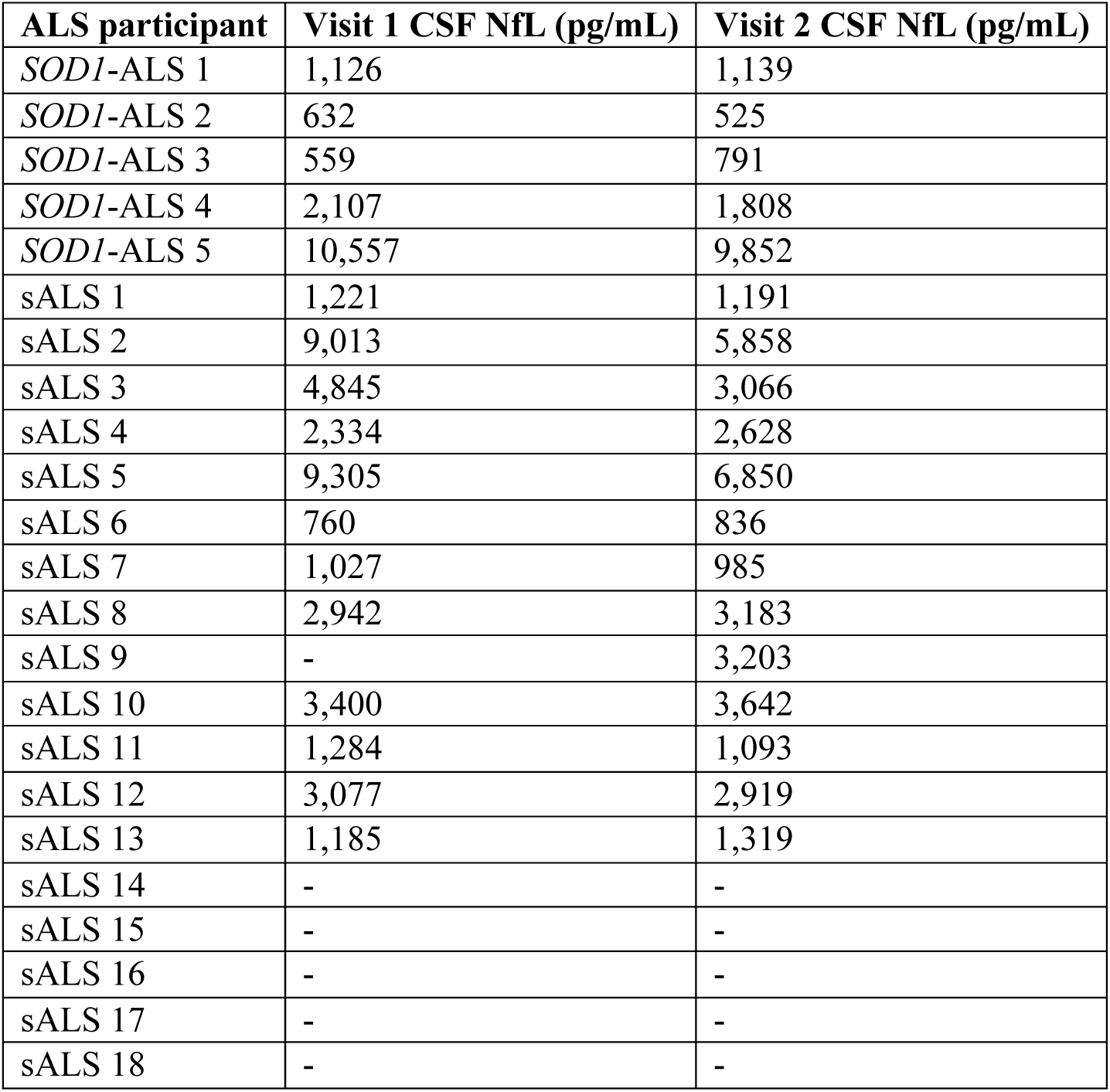
CSF Neurofilament light at initial (visit 1) and subsequent (visit 2) visits from ALS participants. Dashed symbol indicates data was not available.

These results suggest the rate of SOD1 fibril formation increases, as well as the length of SOD1 fibrils, as neuron injury accumulates during ALS progression. A previous study has observed SOD1 and TDP-43 can affect the stability of NfL mRNA transcripts [95]. Thus, further studies measuring misfolded SOD1 via seeding activity in the broader ALS population in conjunction with other CSF ALS biomarkers will enable us to better understand the pathological mechanisms that occur during ALS progression. Examining a larger ALS cohort with varying ALSFRS scores, ALSFRS-R slope decline values, and administered therapy, will enhance our understanding of how CSF SOD1 seeding activity parameters can be influenced by ALS prognosis and therapeutic intervention.

## Conclusions

In conclusion, we probed SOD1 seeding activity in antemortem cerebrospinal fluid from 23 ALS participants (5 *SOD1*-linked ALS and 18 sporadic ALS) as well as 19 healthy controls and 13 disease controls. We examined longitudinal CSF from 18 ALS participants (5 *SOD1*-ALS, 13 sALS). Nine sALS participants were less than 3 years, and 6 sALS participants were less than 2 years from their symptom onset with ALS. Our SOD1 RT-QuIC assay conditions detected SOD1 seeds (mutant or wildtype) in antemortem CSF from *SOD1*-ALS participants with different *SOD1* mutations, and sporadic ALS participants expressing WT SOD1 who had no ALS risk in 10 genes. The sensitive detection of SOD1 seeding activity in antemortem CSF from ALS participants with or without *SOD1* mutations early in their ALS, suggests that WT and mutant SOD1 is misfolded and prion-like relatively early in ALS pathogenesis, and indicates that potential stratification of non-*SOD1* linked ALS participants with SOD1 pathology early in disease is possible and may enable stratification and early intervention for clinical trials. The CSF SOD1 RT-QuIC analyses reported here may be useful for several biomarker context of uses. Testing CSF from a larger cohort of ALS participants, with varying ALSFRS-R scores, ALS-linked mutations, as well as administered therapies, is required to further validate our SOD1 seed amplification assay for prognostic and pharmacodynamic applications.

## Abbreviations

ALS: amyotrophic lateral sclerosis
SOD1: superoxide dismutase 1
sALS: sporadic ALS
fALS: familial ALS
RT-QuIC: real-time quaking induced conversion
SAA: seed amplification assay
CSF: cerebrospinal fluid
ALSFRS-R: amyotrophic lateral sclerosis functional rating scale revised
LC-MS: liquid chromatography mass spectrometry.

## Declarations

### Ethics approval and consent to participate

This study using de-identified human premortem cerebrospinal fluid was conducted in compliance with the Institutional Review Board review of the Weissman Hood Institute at Touro University under protocol MRI1003, and University of British Columbia REB permit (H15-01738) and Annual Renewal number (PAA, H15-01738-A030). This manuscript does not include patient identifiable information, photographs, and consent from patients was not applicable. The article does not contain any studies with animals.

### Consent for Publication

N/A

## Author Contributions

Provided antemortem CSF from ALS participants and controls: CVL, NRC, RH

Performed experiments: MAS, MCF

Analyzed data: MAS, MCF, and MJL

Prepared figures, wrote or edited the manuscript: CLR, HP, MAS, MCF, GYH, NRC, MJL, CVL

## Data Availability

All data produced and analyzed in this study are included in the main article and supplementary information file. The datasets used and analyzed during in this study is available from the corresponding author on reasonable request

## Acknowledgements

We acknowledge Psomagen, who performed Simoa analysis, and Target ALS personnel for providing antemortem CSF from ALS participants with their clinical information. We acknowledge Deborah Cabin, Ph.D., and Renee Reijo-Pera, Ph.D., for editing this manuscript, Byron Caughey, Ph.D., for his discussions regarding RT-QuIC, and Justin Mielke for growing and expressing rSOD1 from *E. coli*. The content is solely the responsibility of the authors and does not necessarily represent the official views of the National Institutes of Health.

